# Cost-effectiveness of CT perfusion for the detection of large vessel occlusion acute ischemic stroke followed by endovascular treatment: A model-based health economic evaluation study

**DOI:** 10.1101/2023.03.16.23287253

**Authors:** Henk van Voorst, Jan W. Hoving, Miou S. Koopman, Jasper D. Daems, Daan Peerlings, Erik Buskens, Hester F. Lingsma, Ludo F.M. Beenen, Hugo W.A.M. de Jong, Olvert A. Berkhemer, Wim H. van Zwam, Yvo B.W.E.M. Roos, Marianne A.A. van Walderveen, Ido van den Wijngaard, Diederik W.J. Dippel, Albert J. Yoo, Bruce C.V. Campbell, Wolfgang G. Kunz, Bart J. Emmer, Charles B.L.M. Majoie, the CLEOPATRA, CONTRAST consortium, MR CLEAN Registry Investigators

**Affiliations:** Department of Radiology and Nuclear Medicine, Amsterdam UMC location University of Amsterdam, Amsterdam, the Netherlands; Department of Biomedical Engineering and Physics, Amsterdam UMC location University of Amsterdam, Amsterdam, the Netherlands; Department of Public Health,Erasmus University Medical Center,Rotterdam,the Netherlands; Department of Neurology,Erasmus University Medical Center, Rotterdam, the Netherlands; Department of Radiology,University Medical Center Utrecht, Utrecht, the Netherlands; Department of Epidemiology, University Medical Center Groningen, University of Groningen, Groningen, the Netherlands; Department of Radiology and Nuclear Medicine,Erasmus University Medical Center, Rotterdam,the Netherlands; Department of Radiology and Nuclear Medicine,Cardiovascular Research Institute Maastricht (CARIM),Maastricht University Medical Center+,Maastricht,the Netherlands; Departement of Neurology, Amsterdam UMC location University of Amsterdam, Amsterdam, the Netherlands; Department of Radiology,Leiden University Medical Center,Leiden,the Netherlands; Department of Neurology,Haaglanden Medical Center,The Hague,the Netherlands; Department of Radiology,Texas Stroke Institute,Dallas-Fort Worth,TX,US; Department of Medicine and Neurology,Melbourne Brain Center,Royal Melbourne Hospital, University of Melbourne,Parkville,Australia; Department of Radiology, University Hospital, LMU Munich, Germany

## Abstract

**Background:** CT perfusion (CTP) has been suggested to increase the rate of large vessel occlusion (LVO) detection in patients suspected of acute ischemic stroke (AIS) if used in addition to a standard diagnostic imaging regime of CT angiography (CTA) and non-contrast CT (NCCT). The aim of this study was to estimate the costs and health effects of additional CTP for endovascular treatment (EVT)-eligible occlusion detection using model-based analyses.

**Methods:** In this Dutch, nationwide retrospective cohort study with model-based health economic evaluation, data from 701 EVT-treated patients with available CTP results were included (January 2018–March 2022; trialregister.nl:NL7974). We compared a cohort undergoing NCCT, CTA, and CTP (NCCT+CTA+CTP) with a generated counterfactual where NCCT and CTA (NCCT+CTA) was used for LVO detection. The NCCT+CTA strategy was simulated using diagnostic accuracy values and EVT effects from the literature. A Markov model was used to simulate 10-year follow-up. We adopted a healthcare payer perspective for costs in euros and health gains in quality-adjusted life years (QALYs). The primary outcome was the net monetary benefit (NMB) at a willingness to pay of € 80,000, secondary outcomes were the difference between LVO detection strategies in QALYs (Δ QALY) and costs (Δ Costs).

**Results:** We included 701 patients (median age:72 IQR:[62-81]) years). Per LVO patient, CTP-based occlusion detection resulted in cost savings (Δ Costs median:€ -2671 IQR:[€ -4721;€ -731]), a health gain (Δ QALY median:0.073 IQR:[0.044;0.104]), and a positive NMB (median:€ 8436 IQR:[5565;11876]) per LVO patient.

**Conclusion:** Adding CTP to NCCT and CTA for EVT-eligible LVO detection resulted in cost savings and health gains.

**Clinical relevance statement:** In recent clinical trials, CTP-based patient selection for endovascular treatment resulted in worse patient outcomes after ischemic stroke. We found that an alternative use of CTP, CTP-based screening for endovascular treatable occlusions is cost-effective.

**Key points:**

- Using CTP to detect an endovascular treatment-eligible occlusions resulted in a health gain and cost savings when considering 10-years of follow-up.
- Depending on the screening costs related to the number of patients needed to image (NNI) with CTP, cost savings could be considerable (Δ Costs NNI=4.3 median:€ -3857 IQR:[€ -5907;€ - 1916]; NNI=8.3 median:€ -2671 IQR:[€ - 4721;€ -731])
- Variations in sensitivity difference due to the use of CTP affect the health gain (Δ QALYs sensitivity difference=baseline median:0.073 IQR:[0.044;0.104]; sensitivity difference=(baseline-4%) median:0.052 IQR:[0.031;0.075]).

## Introduction

Brain tissue perfusion maps derived from computed tomography perfusion (CTP) have been suggested to improve occlusion detection in acute ischemic stroke (AIS) patients if used in addition to CT angiography (CTA) and non-contrast CT (NCCT) (1–4). Although CTP is primarily considered to select patients for endovascular treatment (EVT) (5), screening all suspected AIS patients presenting within six hours after symptom onset with CTP could also enhance the detection of patients with a large vessel occlusion (LVO) EVT; resulting in more patients who benefit from EVT and less missed occlusions (6). However, it remains unclear to what extent the direct costs of screening a large group of patients with CTP for EVT-eligible occlusions result in long-term health gains and cost savings. Several studies found that adding CTP to an imaging regime of non-contrast enhanced CT (NCCT) and CT angiography (CTA) enhances the sensitivity for arterial occlusion detection (1–4). Moreover, the sensitivity gain of adding CTP was between 0-20% (1–4) – depending on the experience of the neuroradiologist and the occlusion location. Since EVT has vastly improved outcomes of AIS patients with a large vessel occlusion (LVO) (6,7), the total quality-adjusted life-years (QALYs) of patients after an AIS can be increased by providing EVT to all eligible patients.

Two previous health economic evaluations concluded that CTP was cost-effective when used jointly for EVT- and intravenous thrombolysis-eligible occlusion detection and to exclude patients with severe ischemia for whom EVT may potentially be harmful (8,9). However, these studies considered deterministic fixed estimates for the value of additional CTP for EVT-eligible LVO detection that do not correspond with recent findings (1,2). In addition, the benefit of CTP-based occlusion detection may be higher for less-experienced physicians compared to experienced neuroradiologists. Furthermore, variations in the proportion of patients with an EVT-eligible occlusion compared to the overall population presenting with AIS symptoms at the emergency department alter the number of patients needed to image (NNI) to detect an LVO and increase the total costs of CTP. Finally, the two studies considered a U.S. perspective that might not apply to other healthcare systems (8,9).

In this study, we aimed to estimate the long-term costs and health effects of adding CTP (NCCT+CTA+CTP) for LVO detection to a standard imaging protocol of NCCT and CTA (NCCT+CTA) in Dutch patients suspected of AIS presenting within six hours after symptom onset at an EVT-capable hospital. Furthermore, we aimed to analyze the effect of variations in the sensitivity of NCCT+CTA+CTP compared to NCCT+CTA based LVO detection, the number of CTPs needed to acquire (NNI) before an LVO was detected, and the benefit of EVT.

## Methods

### Study design

In this study, a cohort of patients that received EVT after NCCT+CTA+CTP based LVO detection with 90-day functional outcome according to the modified Rankin Scale (mRS) was used to simulate long-term mRS and a cohort with 90-day mRS after NCCT+CTA based LVO detection. An EVT-eligible LVO was defined as occlusion of the internal carotid artery-(terminus) (ICA/ICA-T), the M1 or proximal M2 segment of the middle cerebral artery. To generate the NCCT+CTA cohort a percentage of patients was simulated as if they did not receive EVT due to a missed LVO. This percentage of missed LVOs was based on the difference in sensitivity between NCCT+CTA+CTP and NCCT+CTA based LVO detection found in a literature search (**Online Supplement A**), this was referred to as the sensitivity difference (1–4). We did not consider the difference in specificity or positive predictive value due to additional CTP because the relative difference between the imaging strategies would be small and the negative health effects and additional costs of a futile transfer to the angio suite are assumed to be negligible on a population basis. For patients that would not have received EVT under the NCCT+CTA regime, the observed mRS in the included cohort was reduced using available ORs for EVT effect from the literature (**Online Supplement B**) (6,10). To include the additional costs of CTP in the NCCT+CTA+CTP arm, the NNI was used to add additional screening costs per patient with an LVO; for each detected LVO there would be numerous patients without an LVO that were screened with CTP and related costs (**Online Supplement C**).

### Patient level data

We included patients from the Cost-effectiveness of CT perfusion for Patients with Acute Ischemic Stroke (CLEOPATRA) healthcare evaluation study. CLEOPATRA is a healthcare evaluation study using clinical and imaging data from multiple, prospective EVT trials and registries in both the earlier and later time windows (January 2018 – March 2022; trialregister.nl:NL7974) in the Netherlands (11). We only included data from EVT-treated patients with available CTP imaging who presented within six hours after stroke symptom onset. Minor protocol deviations and CTP processing methods are available in **Online Supplement D** and **E**. This study was conducted according to the Helsinki agreement, part of the data has previously been reported (**Online Supplement F**).

### Modeling approach

We simulated 5- and 10-year follow-up using a Markov model with patient-level microsimulations. The Markov model was previously described and validated and consisted of a short-term 90-day post-AIS model followed by a long-term yearly model to simulate functional outcome using the modified Rankin Scale (mRS) (**Figure 1**) (11,12). In the short-term model, we simulated the 90-day mRS of patients that received EVT and those who did not based on NCCT+CTA+CTP or NCCT+CTA based LVO detection. In the long-term model, we simulated yearly mRS deterioration after 90-days based on the probability of stroke recurrence (13) and death (14) inflated with patient-specific Hazard Ratios (HR) (15). Python scripts for the simulations are made publicly available (github.com/henkvanvoorst92/CLEOPATRA).

**Figure 1:**
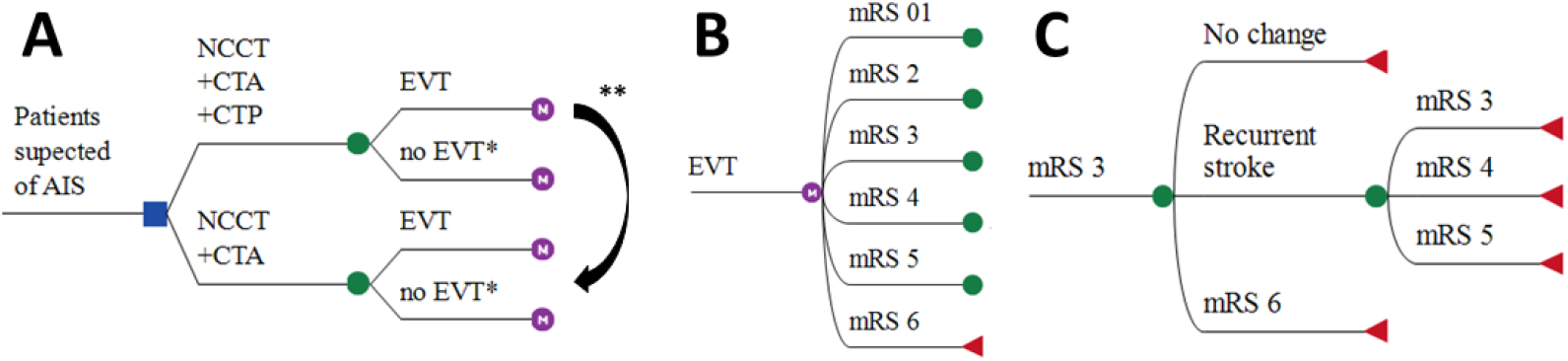
Markov model structure. **1A**: Patients presenting within six hours after stroke symptom onset at an endovascular treatment (EVT)-capable stroke center are subject to one of the following diagnostic imaging protocols for EVT-eligible occlusion detection: (1) non-contrast CT (NCCT), CT angiography (CTA), and CT perfusion (CTP), or (2) NCCT and CTA. No EVT*: In the NCCT+CTA+CTP arm, the number of patients without an EVT-eligible occlusion (no EVT) was computed using the number needed to image (NNI) calculations. Costs of CTP-based screening of non-EVT-eligible occlusions were multiplied with the NNI and added to the overall costs of this simulated arm, in the models’ CTP - no EVT group did not suffer any health consequences and was not further simulated. In the NCCT+CTA arm, the no EVT compromised all patients from the NCCT+CTA+CTP arm in addition to all patients that were missed due to less optimal EVT-eligible occlusion screening. The long-term modified Rankin Scale (mRS) of the missed EVT-eligible occlusion group was further simulated. ** The sensitivity gain due to CTP-based EVT-eligible occlusion detection was used to compute the size of the group of missed EVT-eligible occlusions if a diagnostic imaging protocol consisting of NCCT+CTA was used. **1B**: The 90-day mRS was modeled after EVT or no EVT. **1C**: yearly mRS transitions were modeled based on death and recurrent stroke rates beyond 90 days after stroke. EVT: endovascular treatment. NCCT: non-contrast enhanced CT. CTA: CT angiography. mRS: modified Rankin scale.

### Costs and QALYs

We used mRS over the simulated period to compute cumulative costs from a healthcare payer perspective and QALYs (16). The methodology for acute care and mRS related follow-up QALYs and costs has previously been described (11,16). QALYs were computed per mRS sub-score per year based on 391 patients with 2-year follow-up and available EuroQoL 5D questionnaires (7). Follow-up costs for the first, second, and third year onward per mRS sub-score included: acute setting treatment cost, in-hospital costs, outpatient clinic visits, rehabilitation, formal homecare, and long-term institutionalized costs (7). Acute care costs included NCCT, CTA, CTP, EVT, and IVT if applicable based on reference prices from the institute of Medical Technological Assessment, Rotterdam, the Netherlands (11,16). Acute care costs were increased by 42% to account for hospital overhead costs according to Dutch cost-pricing standards (17). We pre-defined costs per CTP of € 251.40 based on the following assumptions and data: € 129 for the CTP acquisition (17), costs for acute care personnel (€ 94) (17), € 20 for the CTP software license based on expert opinion. EVT costs were € 9,924.50 consisting of material costs and 1.5 hours of personnel costs for (neuro-) interventionist, 1 anesthesiologist, 2 radiology assistants, and 2 anesthesia assistants (11,16). IVT costs of € 950.82 were extracted from m edicijnkosten.nl. Simulations started in 2022. An annual discounting rate of 4% for QALYs and 1.5% for costs was used to compute present values (18). Inflation-based cost adjustments were made using historical and forecasted inflation rates (19,20).

### Outcome measures

Net monetary benefit (NMB: Formula 1) at a willingness to pay (WTP) of € 80.000 per QALY was the primary outcome. Secondary outcomes were the differences in cost (Δ Costs) and quality-adjusted life-years (Δ QALYs) between the intervention (NCCT+CTA+CTP-based LVO detection) and control (NCCT+CTA-based LVO detection) arm. All results were reported as cumulative values over the simulated period with median and interquartile range (IQR) per simulated patient with an LVO.

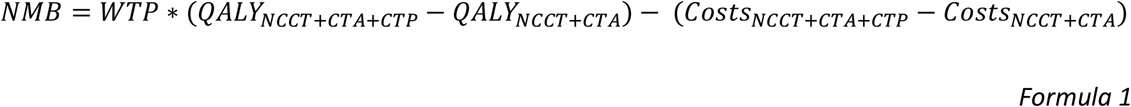

### Baseline and sensitivity analyses

Mean values of the input parameters were used for the baseline simulation considering a 5-year follow-up period. A 10% increase and decrease of all input parameters were used to simulate one-way sensitivity results to assess the outcome variability due to input parameter changes. Probabilistic sensitivity analyses (PSAs) were performed to represent input parameter uncertainty in the outcome measures. For the PSAs, 1,000 cohorts were sampled with replacement from the original data. All model input parameters and distributions are described in **Table 1**.

**Table 1.** Data sources for model input parameter estimation. EVT = endovascular treatment; HR = hazard ratio; IVT = intravenous thrombolysis; mRS = modified Rankin Scale score; OR = odds ratio. QALY: quality-adjusted life year. *Costs are depicted for the reference year 2015. Costs per year included: in-hospital costs, outpatient clinic visits, rehabilitation, formal homecare, and costs for long term institutionalized care. Part of this table has been previously described (11,16).

| Variable | Value | Distribution | Data source |
| --- | --- | --- | --- |
| CTP based sensitivity-gain for ICA occlusions | 2%/4%/6%/8%/10%<br>6% as baseline value | fixed | Olive-Gadea et al. (4) |
| CTP based sensitivity-gain for M1 and M2 occlusions | 12%/14%/16%/18%/20%<br>16% as baseline value | fixed | Becks et al. (1)<br>Bathla et al. (2)<br>Hopyan et al. (3)<br>Olive-Gadea et al. (4) |
| Number needed to image | 4.3/8.3<br>8.3 as baseline value | fixed | Online supplementary C |
| EVT effect | 1.67 (CI:1.21-2.3) (baseline)<br>1.97 (CI:1.51-2.6) (upper)<br>1.37 (CI:0.91-2.0) (lower)<br>2.49 (CI:1.76-3.53) | Log-normal | Berkhemer et al. (10)<br>Goyal et al. (21) |
| Baseline probability of stroke recurrence | Dependent on years after index ischemic stroke | fixed | Pennlert et al. (13) |
| HR recurrent stroke | age and mRS dependent | Log-normal | Pennlert et al. (13) |
| Baseline probability of death | age, gender and year dependent | fixed | Dutch Royal Actuarial Society (14) |
| HR mortality (by mRS: 01/2/3/4/5) | 1.54/2.17/3.18/4.55/6.55 | Log-normal | Hong et al. (15) |
| Inflation rate in % per year | % per year | Fixed value | Central Bureau of Statistics (19) |
| Costs EVT* | €9924.50 | fixed | Van den Berg. (7) |
| Costs IVT* | €950.82 | fixed | Van den Berg. (7) |
| Costs of CTP* | €251.40 | fixed | Estimated in protocol (11) |
| Costs year 1* (by 90 day mRS: 01/2/3/4/5/6) | 33,402(31,930)/52,804(23,571)/82,452(35,333)/<br>112,414(35,786)/96,640(30,463)/21,112(17,350) | Gamma | Van Voorst et al. (12) |
| Costs year 2* (by 18 month mRS: 01/2/3/4/5/6) | 5,934(15,918)/8,543(14,844)/19,235(15,999)/<br>43,193(45,640)/56,425(24,252)/423(3,196) | Gamma | Van Voorst et al. (12) |
| Costs year 2* onward (by 18 month mRS: 01/2/3/4/5/6) | 3,633(9,087)/7,318(13,770)/16,276(11,753)/<br>31,037(19,928)/54,997(24,874)/374(3,118) | Gamma | Van Voorst et al. (12) |
| QALY (by mRS: 01/2/3/4/5/6) | 0.94(0.09)/0.80(0.17)/0.68(0.24)/<br>0.39(0.26)/0.24(0.25)/0(0.01) | Beta | Van Voorst et al. (12) |

*Dedicated PSAs:* We performed dedicated PSAs to analyze the effect of major factors that affect the cost-effectiveness of CTP-based EVT-eligible occlusion detection.

1. To summarize the variations in sensitivity difference between NCCT+CTA+CTP and NCCT+CTA based LVO detection, we performed a systematic search on PubMed combining terms related to CTP, CTA, sensitivity, and stroke with an AND term (**Supplement A**). We defined ranges of sensitivity difference for ICA, M1, and M2 occlusions separately based on the literature (1–4) (**Table 2**). For ICA occlusions, we used a baseline sensitivity difference of 8% and varied values with increments of 2% between 4% and 12% (4). For M1 and M2 occlusions, we used a baseline sensitivity difference of 16% and varied values with increments of 2% between 12% and 20% (2,4).
2. We performed a sensitivity analysis for the treatment effect of EVT that was used to generate the 90-day mRS of patients that would not receive EVT in the NCCT+CTA arm (**Online Supplement B**). The OR for the treatment effect of EVT (OR:1.67 95%CI:[1.21;2.30]) from the MR CLEAN trial (10), the trial with the most conservative EVT benefit (6), was altered with -0.3, +0.3, and +0.82 (6).
3. We used the number needed to image (NNI) to accrue for the costs of all CTPs made; including CTP costs for patients not receiving EVT. We varied the NNI between 4.3 and 8.3, based on previously reported values in the literature and ambulance data from two urban regions in the Netherlands (detailed computations in **Supplement C**). We assumed that 50-60% of all suspected stroke patients admitted to an EVT-capable hospital have an AIS (22) (**Table 3**). Of all AIS patients, 24-46% have an LVO (23).

**Table 2.** Sensitivity difference between NCCT+CTA+CTP and NCCT+CTA for large vessel occlusion detection. CTA: CT angiography, CTP: CT perfusion, ACA: anterior cerebral artery (sub-segments: A1-A3), PCA: posterior cerebral artery (sub-segments: P1-P4), MCA: middle cerebral artery (sub-segments: M1-M4), ICA: internal carotid artery, PICA: posterior inferior cerebellar artery, SCA: superior cerebellar artery, AICA: anterior inferior cerebellar artery. *: Sensitivity-difference was computed as the sensitivity difference between NCCT+CTA+CTP- and NCCT+CTA-based occlusion detection divided by the sensitivity of NCCT+CTA+CTP based occlusion detection. **: Results were reported for multiple raters, we reported this as the rater with the lowest and highest sensitivity-difference (lowest/highest).

| Reference | Definition of occlusion location in study | Number of patients | Number of raters | Sensitivity difference (%)* | Sensitivity with NCCT+CTA (%) | Sensitivity with NCCT+CTA+CTP (%) |
| --- | --- | --- | --- | --- | --- | --- |
| Olive-Gadea et al. (4) | ICA | 22 | 3 | 8.3 | 91.7 | 100 |
|  | M1 | 37 | 3 | 15.6 | 84.4 | 100 |
| Bathla et al. (2) | M2 occlusion | 46 | 2 | 14/18** | 78/76** | 91/93** |
| Hopyan et al. (3) | ICA, ACA, M1-m4, PCA, vertebrobasilar artery, PICA, SCA, other small vessel | 191 | 4 | 20 | 55.4 | 69.5 |
| Becks et al. (1) | Proximal occlusions: ICA, A1, P1, M1, M2, basilar and vertebral artery | 36 | 3 | 0/6** | 89/94** | 89/100** |

**Table 3.**
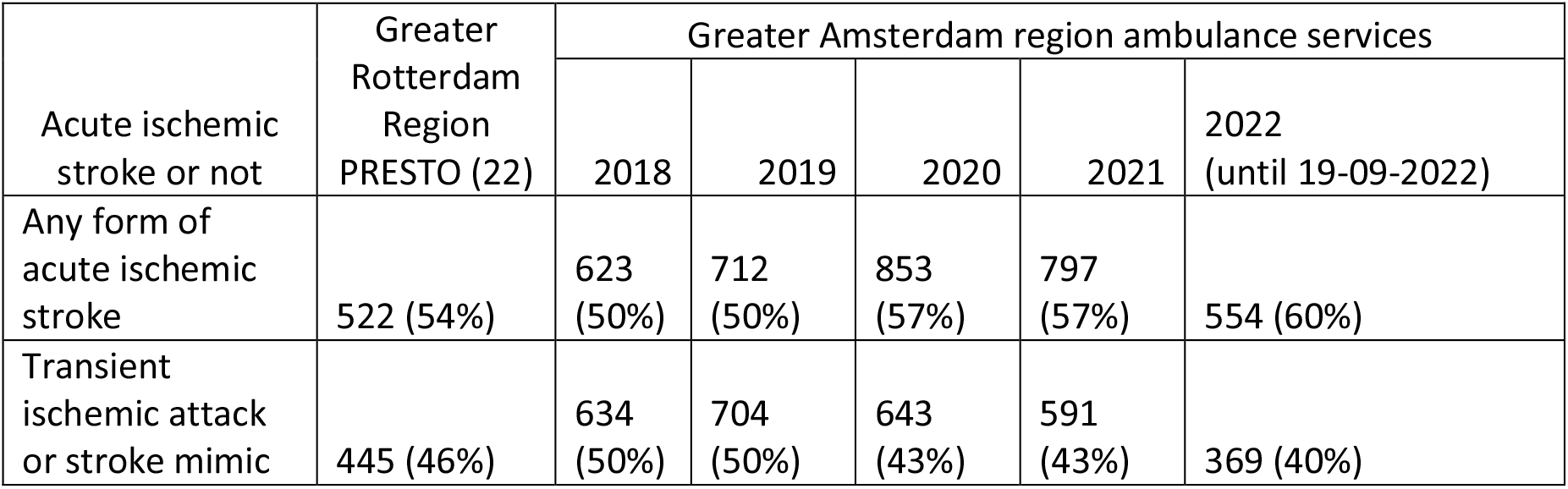
Proportion of acute ischemic stroke in patients suspected for stroke at an emergency ward. Data from the greater Rotterdam are was extracted from the PRESTO trial. Data from the Greater Amsterdam area was obtained from a database on ambulance data and final diagnoses.

| Acute ischemic stroke or not | Greater Rotterdam Region PRESTO (22) | Greater Amsterdam region ambulance services |  |  |  |  |
| --- | --- | --- | --- | --- | --- | --- |
|  |  | 2018 | 2019 | 2020 | 2021 | 2022 (until 19-09-2022) |
| Any form of acute ischemic stroke | 522 (54%) | 623 (50%) | 712 (50%) | 853 (57%) | 797 (57%) | 554 (60%) |
| Transient ischemic attack or stroke mimic | 445 (46%) | 634 (50%) | 704 (50%) | 643 (43%) | 591 (43%) | 369 (40%) |

## Results

### Descriptive statistics

We included 701 (390/701 male, median age 72[IQR:62;81]) of 1122 patients available in the CLEOPATRA database for the simulations. An inclusion flow chart is available in **Figure 2**. Patients were excluded due to an onset of stroke symptoms to groin puncture time beyond 6 hours (n=172), absence of CTP source data (n=91), no accurate CTP results after processing (n=86), double inclusion (n=65), and an unknown occlusion location or a posterior circulation occlusion (n=7). Baseline characteristics for the total population and subgroups based on occlusion location are presented in **Table 4**.

**Figure 2.**
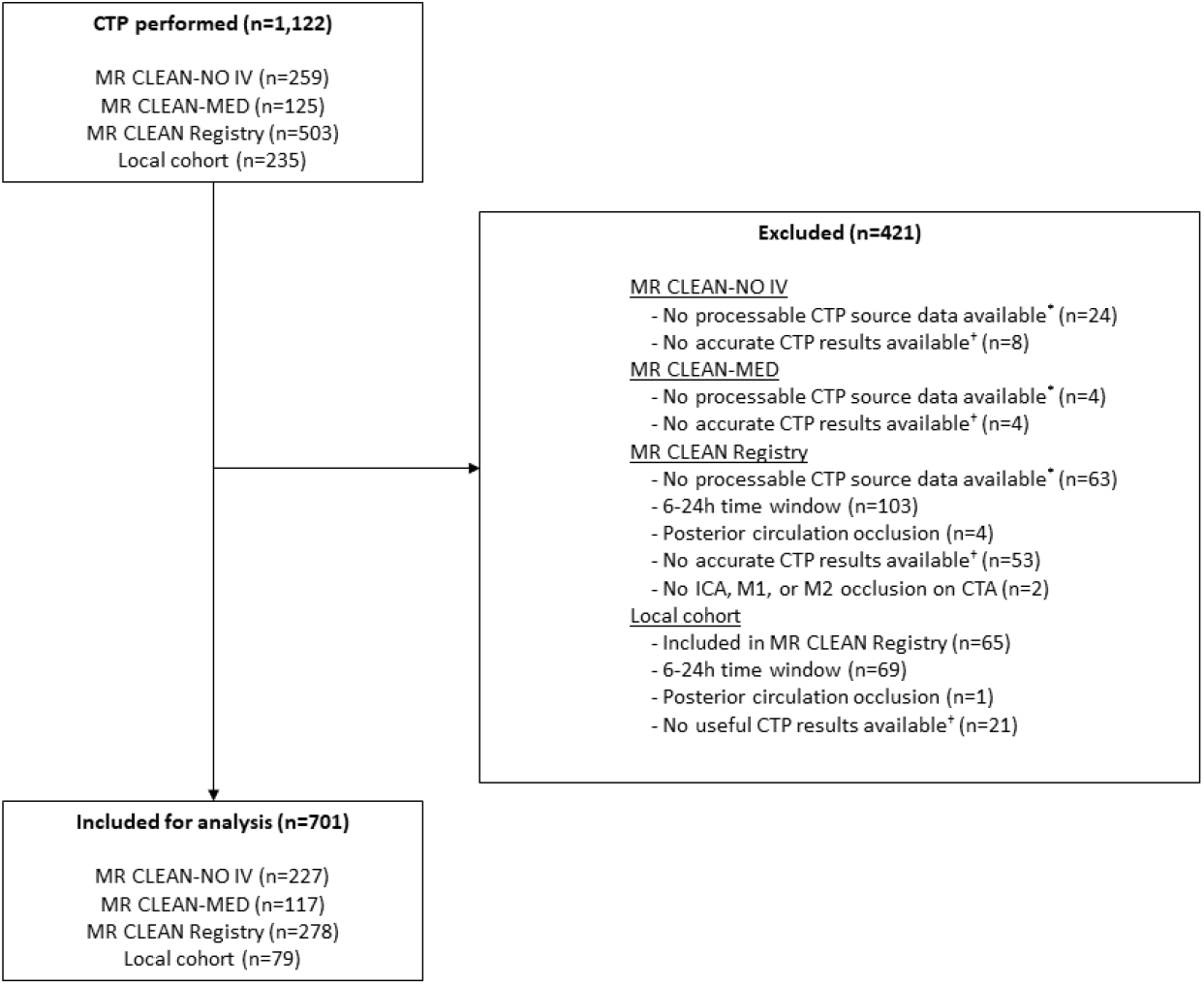
Flowchart of patient selection. ^*^:CTP source data without time information or CTP source data not available due to local storage in the primary stroke center. ^†^Reasons for inaccurate CTP results include severe patient motion, severe curve truncation, no timely contrast arrival or incorrect timing CTP, or severe artifacts in CTP source data. CTP = CT perfusion. ICA: internal carotid artery.

**Table 4.** Descriptive statistics of the included cohort. AIS: acute ischemic stroke. ASPECTS: Alberta Stroke Program Early CT Score. IQR: interquartile range. mRS: modified Rankin Scale. NIHSS: National institute of health stroke. SD: standard deviation. EVT: endovascular treatment.

| Variable | Total population (n=701) | Occlusion location |  |  | p-value |
| --- | --- | --- | --- | --- | --- |
|  |  | ICA or ICA-T (n=168) | M1 (n=367) | M2 (n=166) |  |
| Age (median - IQR) | 72 (62;81) | 70 (59;77) | 72 (62;80) | 73 (65;82) | 0.01 |
| Male sex | 390 (55.6%) | 100 (59.9%) | 200 (54.5%) | 90 (54.2%) | 0.44 |
| Baseline NIHSS (median - IQR) | 15 (10;19) | 17 (13;20) | 16 (10;20) | 11 (7;17) | <1e-5 |
| mRS before AIS |  |  |  |  | 0.53 |
| 0 | 222 (64.9%) | 65 (73.0%) | 117 (61.9%) | 40 (62.5%) |  |
| 1 | 70 (20.5%) | 16 (18.0%) | 39 (20.6%) | 15 (23.4%) |  |
| 2 | 39 (11.4%) | 7 (7.9%) | 25 (13.2%) | 7 (10.9%) |  |
| 3 | 11 (3.2%) | 1 (1.1%) | 8 (4.2%) | 2 (3.1%) |  |
| Baseline ASPECTS (median-IQR) | 9 (8;10) | 8 (7;10) | 9 (8;10) | 10 (9;10) | <1e-5 |
| Baseline collateral score |  |  |  |  | 0.02 |
| 0 | 21 (6.2%) | 10 (11.4%) | 11 (5.9%) | 0 (0%) |  |
| 1 | 90 (26.6%) | 23 (26.1%) | 55 (29.6%) | 12 (18.8%) |  |
| 2 | 160 (47.3%) | 40 (45.5%) | 88 (47.3%) | 32 (50.0%) |  |
| 3 | 67 (19.8%) | 15 (17.0%) | 32 (17.2%) | 20 (31.2%) |  |
| Ischemic core volume in mL (median – IQR) | 13 (5;33) | 18 (6;55) | 11 (5;31) | 13 (5;27) | 0.01 |
| Penumbra volume in mL (mean - SD) | 113 (128) | 145 (247) | 112 (45) | 82 (42) | <1e-3 |
| Core-penumbra mismatch ratio (median - IQR) | 9 (4;19) | 7 (4;19) | 10 (5;21) | 7 (4;15) | <0.01 |
| Onset to groin puncture time in minutes (median - IQR) | 158 (115;230) | 157 (120;226) | 153 (114;228) | 173 (122;238) | 0.45 |
| Door to groin puncture time in minutes (median – IQR) | 67 (50;90) | 68 (51;96) | 64 (49;86) | 70 (53;104) | 0.11 |
| Duration of procedure in minutes (median - IQR) | 48 (31;71) | 60 (40;88) | 42 (29;65) | 45 (31;66) | <1e-3 |
| Intravenous thrombolysis administration | 382 (67.3%) | 86 (66.2%) | 198 (66.7%) | 98 (69.5%) | 0.80 |
| modified Rankin Scale 90-days |  |  |  |  | 0.45 |

| after EVT number (%) |  |  |  |  |
| --- | --- | --- | --- | --- |
| 0 | 47 (7.3%) | 7 (4.6%) | 28 (8.3%) | 12 (7.8%) |
| 1 | 115 (17.8%) | 21 (13.8%) | 61 (18.0%) | 33 (21.4%) |
| 2 | 154 (23.9%) | 35 (23.0%) | 87 (25.7%) | 32 (20.8%) |
| 3 | 69 (10.7%) | 20 (13.2%) | 35 (10.3%) | 14 (9.1%) |
| 4 | 76 (11.8%) | 20 (13.2%) | 43 (12.7%) | 13 (8.4%) |
| 5 | 42 (6.5%) | 11 (7.2%) | 20 (5.9%) | 11 (7.1%) |
| 6 | 142 (22.0%) | 38 (25.0%) | 65 (19.2%) | 39 (25.3%) |

**Table 5:** Probabilistic sensitivity results for different scenarios. From top to bottom, the scenarios become less favorable for additional CTP compared to CTA and NCCT based endovascular treatment-eligible occlusion detection. All scenarios considered an adjusted common OR of 1.67 for the treatment effect of EVT (21). An extended version of this table is available in **Online supplement G**. NNI: number needed to image before one endovascular treatment eligible occlusion is detected. The sensitivity gain of additional CTP represents the percentage-point difference between baseline values of 8% for ICA and 16% for M1 and M2 occlusions. NMB: Net monetary benefit at a willingness to pay of € 80,000 per QALY. Δ : the difference between additional CTP and CTA+NCCT based occlusion detection. Δ Costs: difference in costs; negative values imply a cost saving if CTP is added. Δ QALYs: difference in quality-adjusted life years; negative value implies a gain of health if CTP is added.

| Model settings |  |  | Simulation results |  |  |  |
| --- | --- | --- | --- | --- | --- | --- |
| Years of follow-up | NNI | Sensitivity gain relative to baseline (%-points) | ΔCosts in € median (IQR) | ΔQALY median (IQR) | NMB in € median (IQR) | Fraction of simulations cost-effective (cost-effective/1000) |
| 10 | 4.3 | +8 | -6861<br>(-10016;-3883) | 0.115<br>(0.07;0.163) | 15973<br>(11517;21332) | 0.992 |
| 10 | 4.3 | +4 | -5356<br>(-7972;-2899) | 0.094<br>(0.057;0.134) | 12771<br>(9178;17186) | 0.991 |
| 10 | 4.3 | 0 | -3857<br>(-5907;-1916) | 0.073<br>(0.044;0.104) | 9621<br>(6750;13061) | 0.987 |
| 10 | 4.3 | -4 | -2344<br>(-3807;-960) | 0.052<br>(0.031;0.075) | 6451<br>(4384;8974) | 0.98 |
| 10 | 4.3 | -8 | -859<br>(-1733;-15) | 0.031<br>(0.018;0.045) | 3289<br>(2006;4819) | 0.956 |
| 10 | 8.3 | +8 | -5676<br>(-8830;-2698) | 0.115<br>(0.07;0.163) | 14787<br>(10331;20147) | 0.984 |
| 10 | 8.3 | +4 | -4171<br>(-6786;-1713) | 0.094<br>(0.057;0.134) | 11585<br>(7992;16001) | 0.981 |
| 10 | 8.3 | 0 | -2671<br>(-4721;-731) | 0.073<br>(0.044;0.104) | 8436<br>(5565;11876) | 0.973 |
| 10 | 8.3 | -4 | -1158<br>(-2621;225) | 0.052<br>(0.031;0.075) | 5266<br>(3199;7789) | 0.953 |
| 10 | 8.3 | -8 | 325 (-548;1169) | 0.031<br>(0.018;0.045) | 2103<br>(821;3634) | 0.864 |
| 5 | 4.3 | +8 | -1515<br>(-3130;113) | 0.075<br>(0.051;0.099) | 7451<br>(5051;10223) | 0.98 |
| 5 | 4.3 | +4 | -965<br>(-2314;357) | 0.062<br>(0.042;0.081) | 5839<br>(3863;8106) | 0.977 |
| 5 | 4.3 | 0 | -407<br>(-1476;640) | 0.048<br>(0.032;0.064) | 4200<br>(2640;5965) | 0.969 |
| 5 | 4.3 | -4 | 126<br>(-639;888) | 0.034<br>(0.023;0.046) | 2569<br>(1450;3853) | 0.934 |
| 5 | 4.3 | -8 | 686<br>(209;1152) | 0.021<br>(0.014;0.028) | 935 (260;1752) | 0.817 |
| 5 | 8.3 | +8 | -329<br>(-1945;1298) | 0.075<br>(0.051;0.099) | 6265<br>(3865;9038) | 0.963 |
| 5 | 8.3 | +4 | 220<br>(-1128;1543) | 0.062<br>(0.042;0.081) | 4654<br>(2678;6921) | 0.946 |
| 5 | 8.3 | 0 | 777<br>(-290;1825) | 0.048<br>(0.032;0.064) | 3015<br>(1455;4779) | 0.901 |
| 5 | 8.3 | -4 | 1312<br>(545;2074) | 0.034<br>(0.023;0.046) | 1384<br>(265;2667) | 0.801 |
| 5 | 8.3 | -8 | 1872<br>(1394;2337) | 0.021<br>(0.014;0.028) | -249<br>(-925;566) | 0.416 |

### Baseline model and one-way sensitivity

Using the mean input values, an NNI of 8.3, and 5 years of follow-up for the baseline simulations resulted in a gain of health (Δ QALY:0.049) higher costs (Δ Costs:€ 482), with a positive NMB (€ 3447) when NCCT+CTA+CTP would be used compared to NCCT+CTA for LVO detection. **Figure 3** describes the ten most influential model parameters in a one-way sensitivity analysis. The amount of QALYs attributed to mRS 0-3 were the most important factors affecting the NMB. Costs of CTP and EVT were the most influential cost factors affecting the NMB, long-term follow-up care costs only had a limited effect on the NMB. **Figure S3** (**Online Supplement G**) contains a Kaplan-Maier plot describing the simulated 10-year survival.

**Figure 3.**
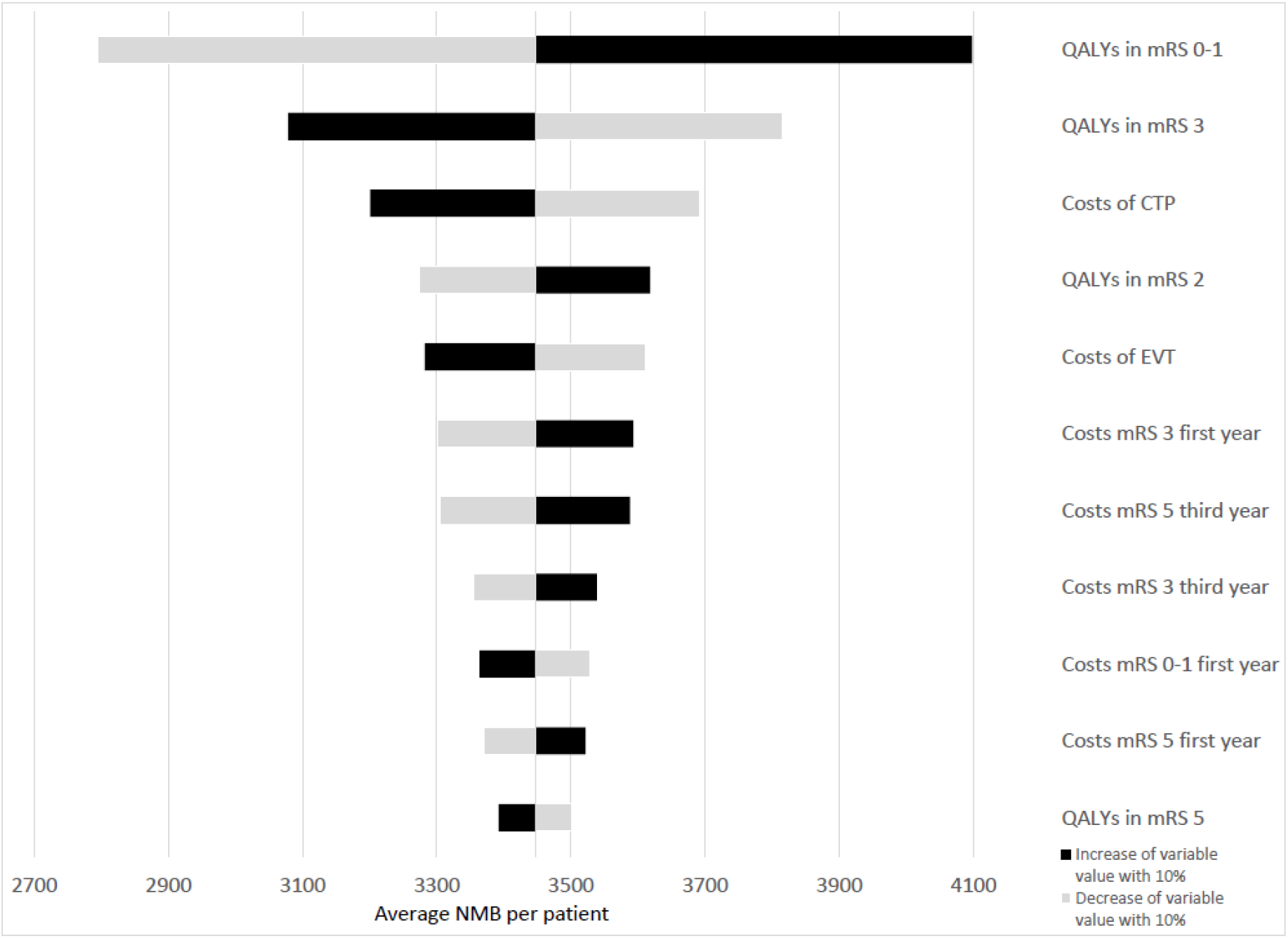
Tornado diagram of the one-way sensitivity analyses. Changes in average NMB compared to the baseline (€ 3,447) are depicted for a 10% increase (black) and decrease (gray) of the ten most influential model input variables. A 5-year horizon was used with an NNI of 8.3 and a baseline sensitivity gain. Variations in NNI, sensitivity gain, and EVT effect were not considered for this analysis. EVT: endovascular treatment, QALY: quality-adjusted life-years, mRS: modified Rankin Scale, CTP: CT perfusion. NMB: net monetary benefit.

### Probabilistic sensitivity analysis

The incremental cost-effectiveness ratio plots per occlusion location and for all occlusion locations together are visualized in **Figure 4**. Considering an NNI of 8.3, the baseline sensitivity difference, and a follow-up horizon of 5 years resulted per EVT-eligible LVO patient in an increase in costs (Δ Costs median:€ 777 IQR:[€ -290;1825]), a health gain (Δ QALY median:0.048 IQR:[0.032;0.064]), and a positive NMB (median:€ 3015 IQR:[€ 1455;€ 4779) when NCCT+CTA+CTP would be used compared to NCCT+CTA for LVO detection. Using similar settings but a 10-year follow-up horizon resulted in a cost-saving (Δ Costs median:€ -2671 IQR:[€ -4721;€ -731]), a health gain (Δ QALY median:0.073 IQR:[0.044;0.104]), and a positive NMB (median:€ 8436 IQR:[€ 5565;€ 11876]).

**Figure 4.**
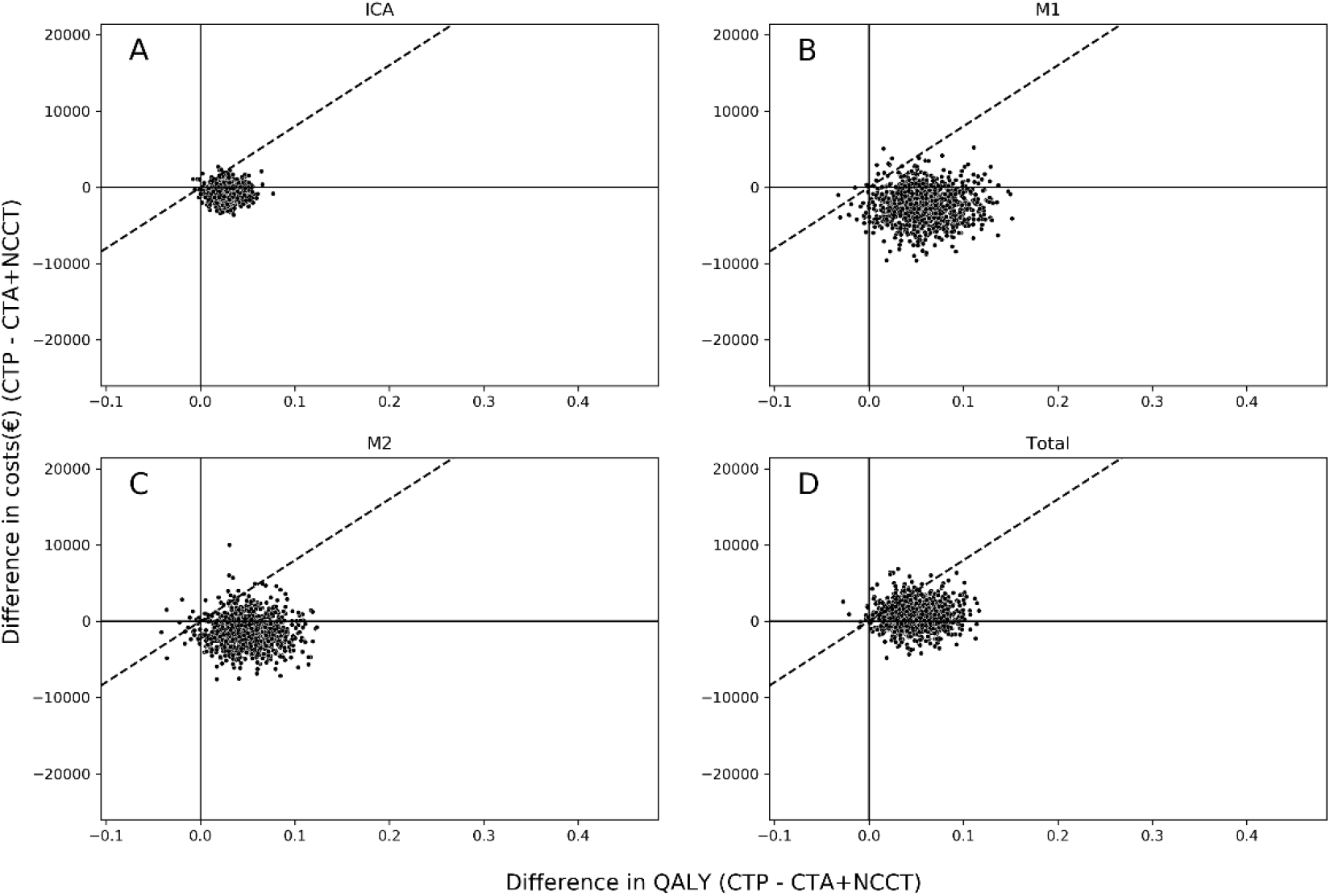
Incremental cost-effectiveness ratio (ICER) plot per occlusion location. Incremental cost-effectiveness ratio plots are presented for (A) ICA, (B) M1, (C) M2, and (D) all simulated patients together. Simulations considered the baseline sensitivity gain per occlusion location, and a 5-year follow-up period. Panels A-C does not include the CTP screening costs, panel D does include the CTP screening costs using the NNI multiplier (NNI=8.3). Positive values represent more costs or QALYs when CTP is included in an imaging protocol consisting of NCCT and CTA for occlusion detection. The dashed diagonal line represents the willingness to pay of € 80,000 per QALY.

### Dedicated sensitivity analyses

Variations in NMB due to the follow-up horizon, NNI, and sensitivity difference are graphically presented in **Figure 5. Table 2** contains the numerical results for Δ QALY, Δ Costs, NMB, and the fraction of the simulations that were cost-effective (below the WTP line; an NMB>0) at a WTP of € 80,000 for varying scenarios. More extensive results for varying model parameters are describe in **Online Supplement G**.

**Figure 5.**
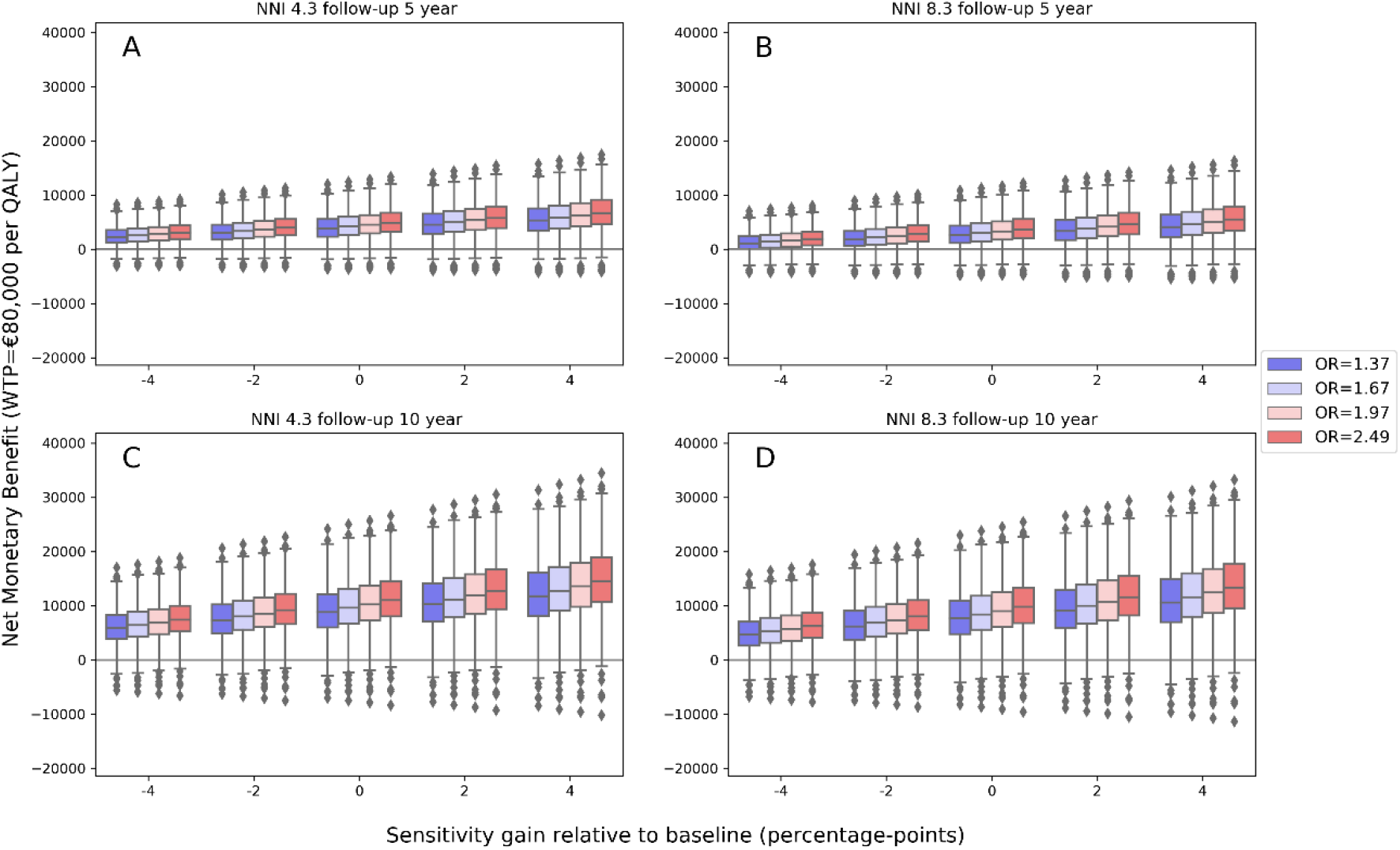
Net monetary benefit for dedicated probabilistic sensitivity analyses. Each panel shows the NMB at a willingness to pay of € 80,000 per quality-adjusted life years (QALY) on the y-axis. A positive net monetary benefit implies that the additional costs of CTP-based screening, EVT, and long-term care costs are lower than the health gain. On the x-axis, the percentage point in sensitivity difference relative to the baseline values of additional CTP (NCCT+CTA+CTP) compared to NCCT+CTA is depicted. The baseline sensitivity gain was 6% for ICA occlusions and 16% for M1 and M2 occlusions. The colors represent the median OR for the treatment effect used for simulations. Panels A-D depicts varying NNI and years of follow-up. A) NNI of 4.3 considering 5-year follow-up. B) NNI of 8.3 considering 5-year follow-up. C) NNI of 4.3 considering 10-year follow-up. D) NNI of 8.3 considering 10-year follow-up. NNI: number of patients needed to image. OR: odds ratio. WTP: willingness to pay. QALY: quality-adjusted life years. NMB: Net monetary benefit.

Between the upper and lower NNI bound additional costs or savings differed (Δ Costs 10-year follow-up: NNI=4.3 median:€ -3857 IQR:[€ -5907;€ -1916] vs. NNI=8.3 median:€ -2671 IQR:[€ - 4721;€ -731]) while the QALYs were the same (Δ QALY median:0.073 IQR:[0.044;0.104]). Variations in sensitivity difference resulted in different health gains (Δ QALYs 10-year follow-up: sensitivity difference=baseline median:0.073 IQR:[0.044;0.104] vs. sensitivity difference=(baseline-4%) median:0.052 IQR:[0.031;0.075] vs. sensitivity difference=(baseline+4%) median:0.094 IQR:[0.057-0.134]). Furthermore, variations in EVT-effect relative to the baseline effect found in the MR CLEAN trial (baseline OR:1.67 95%CI:[1.21;2.30]) resulted in a limited difference in cost savings (NNI=8.3 10-year follow-up: Δ Costs EVT-effect baseline median:€ -2671 IQR:[€ -4721;€ -731]; baseline-0.3 median:€ -2683 IQR:[€ -4715;€ -836]; baseline+0.3 median:€ -2592 IQR:[€ -4622;€ -657]) and more profound variations in health gains (10-year follow-up: Δ QALYs baseline median:0.073 IQR:[0.044;0.104]; baseline-0.3 median:0.062 IQR:[0.034;0.094]; baseline+0.3: 0.082 IQR[0.052;0.113]). NCCT+CTA+CTP based LVO detection would not be cost-effective if we considered 5-year follow-up, an NNI of 8.3, and a sensitivity gain 8% below the baseline (Δ Costs median:€ 1872 IQR:[€ 1394;€ 2337]; Δ QALYs median:0.021 IQR:[0.014;0.028]; NMB median:€ -249 IQR:[€ -925;€ 566]).

## Discussion

In this cohort study with model-based health economic evaluation, we found that adding CTP to an imaging regime of CTA and NCCT for EVT-eligible large vessel occlusion detection followed by endovascular treatment, resulted in a cost savings (Δ Costs median:€ -2671 IQR:[€ -4721;€ -731]), a health gain (Δ QALY median:0.073 IQR:[0.044;0.104]), and a positive net monetary benefit (median:€ 8436 IQR:[5565;11876]) during a 10-year follow-up horizon considering a healthcare payer perspective on a population basis. Costs and health effects per CTP screened patients were small but unevenly distributed; a small percentage of patients that would otherwise not have had the benefits of EVT accounted for all the saved costs and health gains. The number of CTPs performed to detect an endovascular treatment-eligible occlusion, the number needed to image, was the major determinant affecting our results due to the additional costs made per CTP.

Similar to previous research, the use of CTP was found cost-effective (8,9). However, this study provides a more detailed description of factors causing changes in costs and health effects. To interpret these results, we need to consider the currently available evidence. First, NNI estimates might differ from the NNI we used due to population characteristics or inaccuracies in the data we used to estimate the NNI. For example, a higher ischemic stroke prevalence in a population of suspected stroke patients would decrease the NNI and thus result in a lower number of CTPs per detected occlusion and thus lower costs. Second, the benefit in LVO detection sensitivity of additional CTP to a diagnostic workup consisting of NCCT and CTA for AIS screening from previous reader studies varied depending on the physician’s experience (1–4). The value of CTP-based screening might be higher for physicians with less experience in neuroimaging assessments. In this study we reported a wide variety of sensitivity differences as the current evidence comparing NCCT+CTA+CTP with NCCT+CTA based LVO detection is limited by a small sample size and suboptimal research designs (1–4). Third, we used conservative ORs for the EVT treatment effect based on the MR CLEAN trial (12). In recent years, EVT workflows have improved, improving the treatment effect of EVT (24). By underestimating the effect of EVT, we might allow for better generalizability of our findings to settings with different stroke populations or physician’s experience with EVT, such as primary stroke centers or stroke centers in developing countries. Namely, due to the transfer of patients, the onset to groin time increases, negatively affecting the EVT effect (21).

Fourth, the variety of occlusion subtypes in our population might be different from the occlusion subtype distribution in other populations (6). Fifth, costs and health effects due to negative side effects related to CTP were not considered. Allergic reactions and renal insufficiency due to a higher dose of intravenously administered contrast medium may be harmful (25,26). Increased radiation exposure might lead to more malignancies (27). Since these side effects occur in a very small portion of the population, often have limited long-term effects, or occur in the far future, we expect this to only have a minor effect on the costs and health effects in this study. Sixth, current trials studying EVT for more distal occlusions might result in even higher health gains due to CTP-based occlusion detection (28,29).

Our study has limitations. This is a model-based study using a cohort of only patients receiving EVT, our findings might deviate in prospectively based trial cost-effectiveness analyses. Specifically, we did not have data on long-term functional outcome follow-up or micro-level-estimated costs. Furthermore, we used cost data from a healthcare payer perspective and did not consider indirect healthcare spending often included when a societal perspective for a cost-effectiveness study is used. Adopting a societal perspective for this study would lead to a higher cost-saving benefit due to CTP.

Future research should analyze alternative diagnostic imaging strategies compared to the direct use of NCCT, CTA, and CTP used in this study. For example, CTP after an inconclusive CTA and NCCT examination, or the use of population characteristics and clinical symptoms to select patients where CTP might be valuable for LVO detection, could be considered as alternatives. Furthermore, extending the findings of this study to patients presenting after six hours of symptom onset needs to be studied further. Since we only considered EVT-eligible LVOs, future studies should consider the value of CTP for detecting EVT-eligible distal vessel occlusions or posterior circulation occlusions – which are generally more difficult to detect. Finally, future work should aim to determine stroke subtype statistics to compute the NNI and assess determinants for varying sensitivity differences due to CTP in a large reader study.

## Conclusion

In this model-based health economic evaluation study, adding CTP to an imaging regime of NCCT and CTA for EVT-eligible large vessel occlusion detection within 6 hours after symptom onset was cost-effective due to a health gain for limited additional costs to a cost saving when considering a 10-year follow-up horizon. Health gains and cost savings might be more favorable for a lower number needed to image, a longer follow-up horizon, and a higher sensitivity difference of additional CTP compared to NCCT and CTA based large vessel occlusion detection.

## Data Availability

Data will be made available upon reasonable request. Specific access can be granted through a secure environment.

https://www.contrast-consortium.nl/

https://www.mrclean-trial.org/

## Abbreviations

AIS: Acute ischemic stroke
EVT: Endovascular treatment
ICA: internal carotid artery
ICA-T: terminus of ICA
ICER: Incremental cost-effectiveness ratio
IQR: Interquartile range
NMB: Net monetary benefit
NNI: Number needed to image
PSA: Probabilistic sensitivity analysis
QALY: Quality-adjusted life year

## Notes

### Competing Interest Statement

Disclosures and conflicts of interest
BJE reports grants from LtC (ZonMW and TKI-PPP of Health Holland). WHvZ reports personal fees from Codman and from Stryker. DWJD report grants from the Dutch Heart Foundation, AngioCare, Medtronic/Covidien/EV3, MEDAC/LAMEPRO, Penumbra, Top Medical/Concentric, Stryker, and Cerenovus; consultation fees from Stryker, Bracco Imaging, and Servier, received by the Erasmus University Medical Centre outside this project. CBLMM reports grants from TWIN, during the conduct of the study and grants from CVON/Dutch Heart Foundation, European Commission, Dutch Health Evaluation Program, and from Stryker outside this project (paid to institution) and is shareholder of Nicolab. AJY reports Research grants from Medtronic, Cerenovus, Penumbra, Stryker, and Genentech. Consultant for Penumbra, Cerenovus, Nicolab, Philips, Vesalio, Zoll Circulation, and NIH/NINDS. YR is a shareholderof Nicolab. Equity interests in Insera Therapeutics and Nicolab. All other contributors report no conflicts of interest.

### Clinical Protocols

https://journals-sagepub-com.stanford.idm.oclc.org/doi/full/10.1177/23969873221092535

### Funding Statement

Funding statement
The CLEOPATRA healthcare evaluation study was funded by Leading the Change (LtC). LtC is financed by Zorgverzekeraars Nederland (ZN) and supports various healthcare evaluations in the Netherlands as part of the Healthcare Evaluation Netherlands project. LtC was not involved in the study design, monitoring, data collection, statistical analyses, interpretation of results, or manuscript writing, but the progress of the study was continuously monitored by LtC. The MR CLEAN-NO IV trial (ISRCTN80619088) and MR CLEAN-MED trial (ISRCTN76741621) were part of the Collaboration for New Treatments of Acute Stroke (CONTRAST) consortium. The CONTRAST consortium acknowledges the support from the Netherlands Cardiovascular Research Initiative, an initiative of the Dutch Heart Foundation (CVON2015-01: CONTRAST), and from the Brain Foundation Netherlands (HA2015.01.06). The collaboration project is additionally financed by the Ministry of Economic Affairs by means of the PPP Allowance made available by the Top Sector Life Sciences & Health to stimulate public-private partnerships (LSHM17016). This work was funded in part through unrestricted funding by Stryker, Medtronic and Cerenovus. The funding sources were not involved in study design, monitoring, data collection, statistical analyses, interpretation of results, or manuscript writing.

### Author Declarations

Patient inclusion was in adherence with the declaration of Helsinki and was subject to an ethical board review. The CLEOPATRA study protocol has been reviewed by the Amsterdam UMC ethical review board and was waived for informed consent (Amsterdam UMC reference: W19_281#19.334). Retrospective, large-scale, observational studies do not fall under the Medical Research Involving Human Subjects Act (WMO). For patients included in the MR CLEAN-NO IV (ISRCTN80619088, registered 31 October 2017), and MR CLEAN MED (ISRCTN76741621, registered 7 December 2017) trials informed consent has been received previously. The ethical review board of the Erasmus MC has waived the requirement for informed consent for patients included in the MR CLEAN Registry (internal reference Erasmus MC: MEC-2014-235, 27 August 2014).

