## Supplementary material for "Cost-effectiveness of CT perfusion for the detection of large vessel occlusion acute ischemic stroke followed by endovascular treatment: A model-based health economic evaluation study": Online Supplement

**Supplement A: Sensitivity difference between NCCT+CTA+CTP and NCCT+CTA based LVO detection**

**Supplement B: Methods for computing the mRS of a virtual control arm**

**Supplement C: Estimation of the number needed to image**

**Supplement D: Protocol deviations**

**Supplement E: CTP acquisition, post-processing, and quality assessment**

**Supplement F: Institutional review board approval and previous use of data**

**Supplement G: Supplementary Results**

Figure S1: Kaplan Maier plot of simulated baseline cohort

Figure S2: Differences in quality-adjusted life years for deditcated probabilistic sensitivity analyses.

Figure S3: Difference in costs for dedicated probabilistic sensitivity analyses

Table S1: Extended results for different scenarios

**Supplement A: Sensitivity difference between NCCT+CTA+CTP and NCCT+CTA based LVO detection**

We performed a systematic pubmed-search (29-08-2022) combining terms related to CTP, CTA, sensitivity, and stroke with an AND term. Below are the terms combined with and OR term per subheading, we considered the terms if they were available in the title or abstract. A * represents any letters.

**CTP: CT perfusion [tiab] OR CTP [tiab] OR perfusion [tiab]**

**CTA: CT angio* [tiab] OR CTA [tiab]**

**Outcome: Sensitiv* [tiab] OR AUC [tiab]**

**Stroke: (stroke [tiab] AND ischemic [tiab]) OR (ischemic stroke [tiab] OR infarct [tiab] OR ischaemic stroke [tiab]**

The search resulted in 108 articles of which 4 articles reported the sensitivity of additional CTP compared to CTA and NCCT for occlusion detection (1–4). Supplementary **Table S1** summarizes the results and the sensitivity-difference computed using NCCT+CTA+CTP as reference.

**Supplement B: Methods for computing the mRS of a virtual control arm**

To compute the effectiveness of CTP based EVT patient selection a control arm that did not receive EVT was required (noEVT). Since we did not use randomized data, we computed the probability of specific mRS scores of a virtual control arm based on available treatment effect and treatment effect modification odds ratios. Sensitivity analyses were performed for varying values of these odds ratios. For the control arm mRS simulation we need to have the mRS and baseline characteristics an EVT treated patient, a distribution of a population treated with EVT and a control arm (we used the MR CLEAN trial data), and the OR for treatment effect ($OR(mRS|EVT) = 1.67$) and treatment effect modification ($OR(mRS|EVT * ICV) = 0.98$ per 10 mL) related to mRS. We assume in the example below a patient with mRS 2 after EVT with a core volume at baseline of 60 mL. The following steps were taken:

Based on the ORs we can compute the combined OR for a specific patient that received EVT as if she did not receive EVT (inverse of the OR: formula 1 and 2). Taking the natural log of an OR gives us the absolute mRS shift (formula 3). We can use this absolute shift to compute the effect on outcome of ICV ($ICV(10mL)$) and add this with the effect of noEVT to compute the combined OR (formula 4).

$$OR\ (mRS|noEVT) = \frac{1}{OR(mRS|EVT)}$$

Formula 1

$$OR\ (mRS|noEVT * ICV) = \frac{1}{OR(mRS|EVT * ICV)}$$

Formula 2

$$absolute\ mRS\ shift = \ln\ (OR)$$

Formula 3

$$OR\ (mRS\,|\,noEVT\ \&\ ICV) = e^{\ln\left(OR\ (mRS|noEVT)\right)*1+\ln\ (OR\ (mRS|noEVT\ *\ ICV)*ICV(per\ 10mL)}$$

Formula 4

For the next steps we use the $OR(mRS|EVT)$ (=1.67), the total number of patients with EVT in the literature source for treatment effect (MR CLEAN trial n(EVT)=233), the number of patient with EVT achieving mRS≤2 $n(mRS \leq 2\ \&\ EVT)$ (=76), the probability of the patients' mRS given EVT $p(mRS \leq 2|EVT)$ (=0.33), the total number of patients in the control arm $n(noEVT)$ (=267), the number of patient in the control arm achieving mRS≤2 $n(mRS \leq 2\ \&\ noEVT)$ (=51), and the probability mRS≤2 in the control arm $p(mRS \leq 2|noEVT)$ (=0.19). We use the probability and OR for mRS≤2 to get stable estimates irrespective of the simulated patients' mRS; low sample sizes for mRS 0 and unreasonably large for mRS 6 resulting in inaccurate outcome simulations.

Based on the standard formulas to compute the OR (formulas 5-7) and the new computed combined OR based on literature OR and patient observations from formula 4, it is possible to derive formula 8 for computing $Odds(mRS \leq 2|noEVT)$.

$$p(mRS \leq 2|EVT) = \frac{n(mRS \leq 2\ \&\ EVT)}{n\ total\ EVT}$$

Formula 5

$$Odds(mRS \leq 2|EVT) = \frac{p(mRS \leq 2|EVT)}{1 - p(mRS \leq 2|EVT)}$$

Formula 6

$$OR(mRS \leq 2|EVT) = \frac{Odds(mRS \leq 2|EVT)}{Odds(mRS \leq 2|noEVT)}$$

Formula 7

$$Odds(mRS \leq 2|noEVT) = Odds(mRS \leq 2|EVT) * OR(mRS \leq 2|EVT)$$

Formula 8

Finally, after combining formulas 5-8 using a patient specific OR from formula 4 you can compute the virtual probability of mRS shift in the control arm (formula 9 using formulas 5-8). The results per formula for our example are described in the section below formula 9. After solving formula 9, the value of $p(mRS \leq 2|noEVT)$ is used to compute a 1 point shift. In the example below, if a patient has an mRS of 3 with EVT, if no EVT was given she has a probability of 0.25 for an mRS of 3 and 0.75 for an mRS of 4.

$$p(mRS \leq 2|noEVT) = (1 - p(mRS \leq 2|noEVT)) * \ Odds(mRS \leq 2|noEVT)$$

Formula 9

**Example results:**

$$OR\ (mRS|noEVT) = \frac{1}{1.67} = 0.60$$

Formula 1

$$OR\ (mRS|noEVT * ICV) = \frac{1}{0.98} = 1.02$$

Formula 2

$$OR\ \left(mRS\ \middle|\ noEVT\ \&\ ICV\right) = e^{\ln(0.60)*1+\ln\ (1.02)*6} = 0.67$$

Formula 4

$$p(mRS \leq 2|EVT) = \frac{76}{233} = 0.33$$

Formula 5

$$Odds(mRS \leq 2|EVT) = \frac{0.33}{1 - 0.33} = 0.49$$

Formula 6

$$Odds(mRS \leq 2|noEVT) = \ 0.49 * 0.60 = 0.33$$

Formula 8

$$p(mRS \leq 2|noEVT) = (1 - p(mRS \leq 2|noEVT)) * 0.33$$

Formula 9

$$1.33 * p(mRS \leq 2|noEVT) = 0.33$$

$$p(mRS \leq 2|noEVT) = \frac{0.33}{1.33} = 0.25$$

Solving formula 9

The probability of mRS≤2 for the no EVT arm could deviate from observed values in the MR CLEAN trial. This could be due to inaccuracies in the proportional odds assumption or due to different patient characteristics in the EVT arm compared to the no EVT arm.

**Supplement C: Number needed to image estimation**

*Number needed to image*

The number needed to image (NNI) to detect an LVO (EVT-eligible occlusion) was derived from literature sources and ambulance data from two larger urban regions. In the NCCT+CTA+CTP arm all LVOs were detected and received EVT, in the NCCT+CTA arm a percentage of patients was missed and thus did not receive EVT. We added additional CTP screening costs to patients in the NCCT+CTA+CTP arm to accrue for other patients that had a CTP but no EVT-eligible occlusion. The number needed to image (NNI) before detecting an EVT-eligible occlusion was used in our study to add additional costs to the CTP arm. The following information and assumptions were used to compute the NNI:

1. Of all patients suspected to have a stroke at the emergency department 50-60% have an AIS (either LVO or non-LVO) (see **Table 3** in the main manuscript). We used the lower bound value of 50%.
2. Of all patients with an acute ischemic stroke 24-46% have a large vessel occlusion (ICA or MCA) (5).

The NNI computation following from points 1-2 above:

1. Approximately 50% have any form of AIS. Thus, the NNI to screen with CTP to detect an acute ischemic stroke is 100/50 = 2
2. To detect a large vessel occlusion between 2*100/24=8.3 and 2*100/46=4.3 patients are scanned with a CTP.

The NNI with CTP to detect any EVT-eligible occlusion is between 4.3 and 8.3.

**Supplement D: Protocol deviations**

The presented work is based on a previously published protocol paper (6) and the corresponding protocol online (trial registration number: NL7974; trialsearch.who.int/Trial2.aspx?TrialID=NL7974). Several deviations to the protocol are in the presented work, in the section below we describe our reasons to do so.

**Research question**

This study did not concern the main research question of the protocol paper. This study focused on occlusion detection using CTP.

**Default odds ratio EVT effect**

The protocol reported a default scenario adjusted common OR of 1.86 (CI:1.34-2.59) for EVT effect. This was a deviation of the original MR CLEAN trial describing an adjusted common OR of 1.67 (CI:1.21-2.3). Since the scenario with an OR of 1.67 for EVT would be more beneficial for CTP selection, and our findings were negative regardless, we used an OR of 1.67 value. In additional sensitivity analyses (table 1), the value of 1.97 was also considered.

**Probabilistic simulations**

We altered the number of resamples from 10,000 to 1,000 for computational purposes. We altered the sampled cohort size of our initial simulation to the original cohort size (701) instead of 100. When using cohorts of 100 patients the cohort frequently did not include large ICV making it impossible to analyze the effect of patient selection in terms of ICER due to a very low or zero QALY difference.

**Supplement E: CTP acquisition, post-processing, and quality assessment**

CTP images were acquired according to local protocols per site. CTP data were centrally post-processed using syngo.via CT Neuro Perfusion (version VB40, Siemens Healthineers, Forchheim,

Germany). The ICV was estimated as CBV <1.2 mL/100 mL and critically hypoperfused tissue volume (including penumbra) as CBF <27 mL/100 mL/min. A default smoothing filter was applied (7). Visual quality assessment of the post-processed data was performed by two experienced neuroradiologists (>10 and >15 years of experience). The penumbra was defined as critically hypoperfused volume minus ischemic core volume. The mismatch ratio was calculated as the critically hypoperfused volume divided by the ischemic core volume. Clinical variables were collected for simulation purposes, descriptive statistics, and subgroup analyses.

**Supplement F: Institutional review board approval and previous use of data**

*Ethical review board approval*

Patient inclusion was in adherence with the declaration of Helsinki and was subject to an ethical board review. The CLEOPATRA study protocol has been reviewed by the ANONYMOUS ethical review board and was waived for informed consent (ANONYMOUS reference: W19_281#19.334). Retrospective, large scale, observational studies do not fall under the Medical Research Involving Human Subjects Act (WMO). For patients included in the MR CLEAN-NO IV (ISRCTN80619088, registered 31 October 2017), and MR CLEAN MED (ISRCTN76741621, registered 7 December 2017) trials informed consent has been received previously. The ethical review board of the Erasmus MC has waived the requirement for informed consent for patients included in the MR CLEAN Registry (internal reference Erasmus MC: MEC-2014-235, 27 August 2014).

*Previous use of data*

Of all 701 patients considered in this study, data from 622 have previously been reported. Patients were included in the MR CLEAN NO IV trial (n=227) , MR CLEAN MED trial (n=117), and MR CLEAN Registry (n=278). All included patients were selected as a subset of the overall MR CLEAN NO IV, MR CLEAN MED, and MR CLEAN Registry populations. We used patients included in these studies to extract population statistics (age, sex) and functional outcomes that, combined with literature, were used to simulate long term functional outcome, costs, and QALYs. Based on simulations we describe the effect on costs and QALYs of various scenarios for screening patients with CTP. Current other studies did not have related research aims or hypotheses. We selected these patients based on the presence of CTP before EVT to describe a realistic cohort of patients that receive CTP.

Patients included in the MR CLEAN NO IV (n=227) (8) and MR CLEAN MED (n=117) (9) have been studied in a randomized controlled trial setting for the effect of intravenous alteplase (0.9mg/kg) before EVT and the effect of heparin and aspirin before endovascular treatment respectively. Functional outcome and population characteristics of the patients included MR CLEAN Registry (n=278) have been reported before (10).

**Supplement G: Supplementary Results**

**Supplementary Figure S1** includes a Kaplan Maier plot of the baseline simulation. In **Supplementary Table S1** and **Supplementary Figures S2-3** the results for 5- and 10- year follow-up, more number needed to image (NNI) and sensitivity-gain values, and lower endovascular treatment effect are presented.

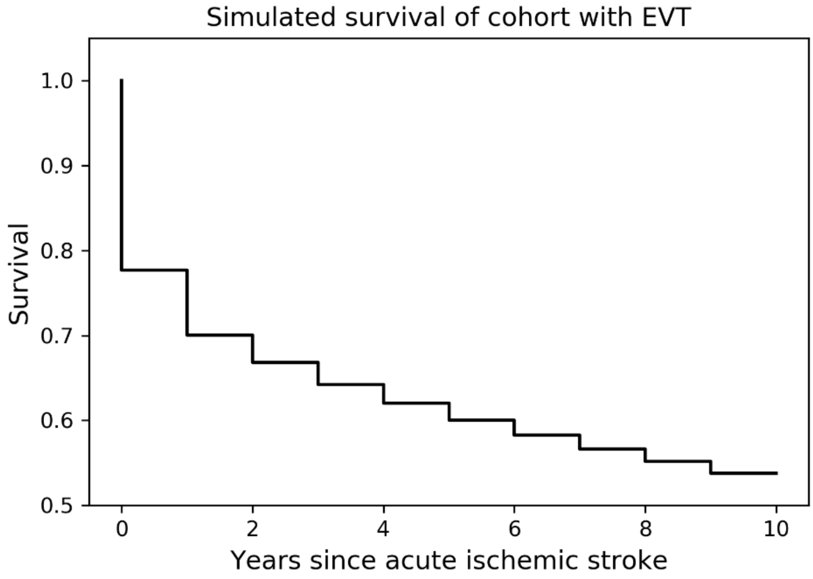


**Supplementary figure S1. Survival function of simulated baseline cohort.** Kaplan-Maier plot of the simulated survival of the included cohort that received EVT using the baseline simulation model inputs. This simulation method has previously been reported and validated (15). EVT: endovascular treatment

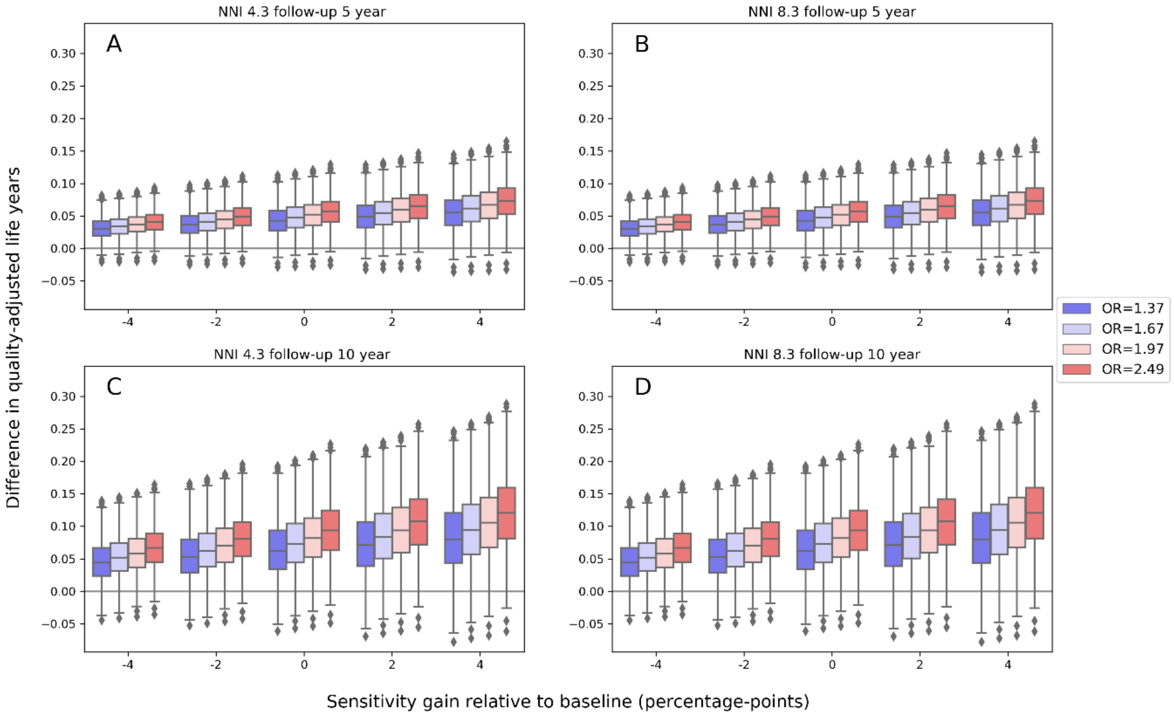


**Supplementary Figure S2. Differences in quality-adjusted life years for deditcated probabilistic sensitivity analyses**. Differences in quality-adjusted by sensitivity gain, NNI, the OR the EVT effect, years of follow-up. NNI: number of patients needed to image. OR: odds ratio. EVT: endovascular treatment.

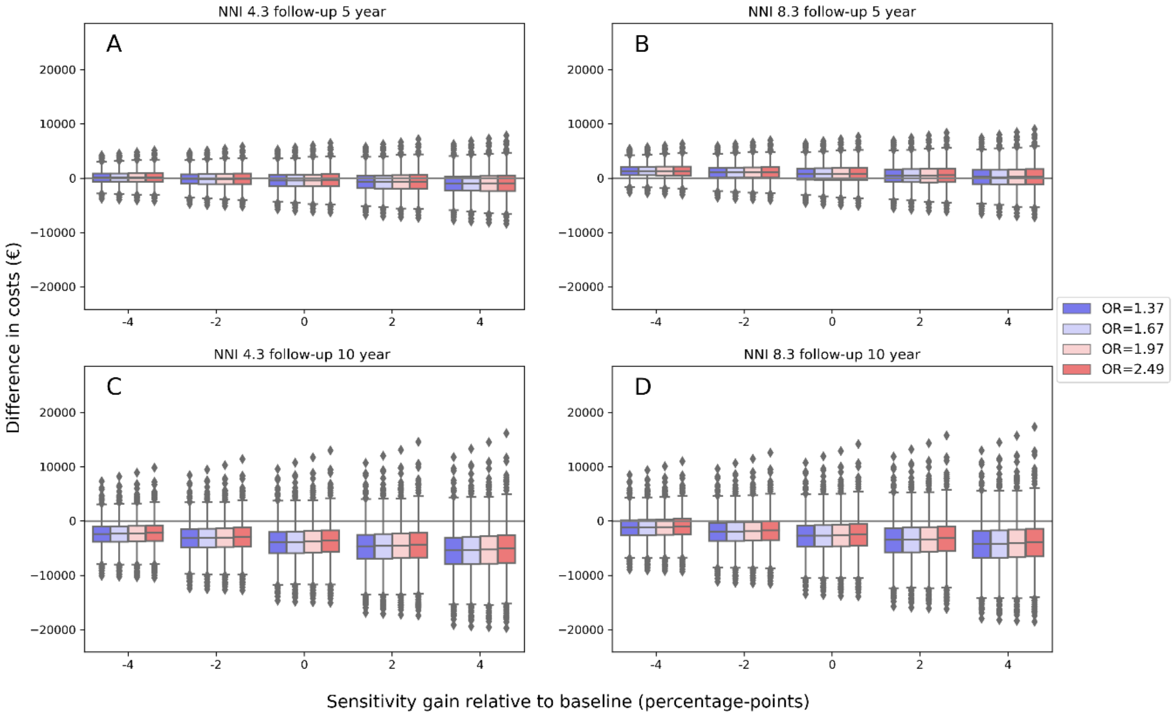


**Supplementary Figure S3 Difference in costs for dedicated probabilistic sensitivity analyses**.

Differences in costs by by sensitivity gain, NNI, the OR the EVT effect, years of follow-up. NNI: number of patients needed to image. OR: odds ratio. EVT: endovascular treatment.

| Model settings | | | | Simulation results | | | |
|---|---|---|---|---|---|---|---|
| Years of follow-up | NNI | EVT effect OR | Sensitivity-gain relative to baseline | ΔCosts in € median (IQR) | ΔQALY median (IQR) | NMB median (IQR) | Percentage of simulation cost effective |
| 10 | 4.3 | 1.97 | 8 | -6739 (-9926;-3708) | 0.13 (0.083;0.176) | 16973 (12352;22221) | 0.993 |
| 10 | 4.3 | 1.97 | 4 | -5271 (-7853;-2769) | 0.106 (0.067;0.145) | 13627 (9819;17990) | 0.991 |
| 10 | 4.3 | 1.97 | 0 | -3777 (-5808;-1843) | 0.082 (0.052;0.113) | 10237 (7315;13693) | 0.989 |
| 10 | 4.3 | 1.97 | -4 | -2302 (-3740;-887) | 0.058 (0.037;0.081) | 6873 (4770;9387) | 0.982 |
| 10 | 4.3 | 1.97 | -8 | -819 (-1695;33) | 0.035 (0.022;0.049) | 3576 (2264;5094) | 0.965 |
| 10 | 4.3 | 1.67 | 8 | -6861 (-10016;-3883) | 0.115 (0.07;0.163) | 15973 (11517;21332) | 0.992 |
| 10 | 4.3 | 1.67 | 4 | -5356 (-7972;-2899) | 0.094 (0.057;0.134) | 12771 (9178;17186) | 0.991 |
| 10 | 4.3 | 1.67 | 0 | -3857 (-5907;-1916) | 0.073 (0.044;0.104) | 9621 (6750;13061) | 0.987 |
| 10 | 4.3 | 1.67 | -4 | -2344 (-3807;-960) | 0.052 (0.031;0.075) | 6451 (4384;8974) | 0.98 |
| 10 | 4.3 | 1.67 | -8 | -859 (-1733;-15) | 0.031 (0.018;0.045) | 3289 (2006;4819) | 0.956 |
| 10 | 4.3 | 1.37 | 8 | -6914 (-10054;-4022) | 0.098 (0.054;0.147) | 14766 (10345;20022) | 0.99 |
| 10 | 4.3 | 1.37 | 4 | -5406 (-7969;-3023) | 0.08 (0.044;0.121) | 11795 (8189;16124) | 0.987 |
| 10 | 4.3 | 1.37 | 0 | -3869 (-5900;-2021) | 0.062 (0.034;0.094) | 8870 (5993;12187) | 0.983 |
| 10 | 4.3 | 1.37 | -4 | -2383 (-3813;-1028) | 0.044 (0.024;0.067) | 5917 (3856;8280) | 0.975 |
| 10 | 4.3 | 1.37 | -8 | -875 (-1743;-73) | 0.026 (0.014;0.04) | 2963 (1687;4447) | 0.947 |
| 10 | 8.3 | 1.97 | 8 | -5554 (-8740;-2522) | 0.13 (0.083;0.176) | 15787 (11166;21036) | 0.989 |
| 10 | 8.3 | 1.97 | 4 | -4085 (-6668;-1583) | 0.106 (0.067;0.145) | 12442 (8633;16805) | 0.983 |
| 10 | 8.3 | 1.97 | 0 | -2592 (-4622;-657) | 0.082 (0.052;0.113) | 9052 (6129;12507) | 0.978 |
| 10 | 8.3 | 1.97 | -4 | -1117 (-2555;298) | 0.058 (0.037;0.081) | 5688 (3585;8202) | 0.964 |
| 10 | 8.3 | 1.97 | -8 | 366 (-510;1219) | 0.035 (0.022;0.049) | 2391 (1079;3909) | 0.882 |
| 10 | 8.3 | 1.67 | 8 | -5676 (-8830;-2698) | 0.115 (0.07;0.163) | 14787 (10331;20147) | 0.984 |
| 10 | 8.3 | 1.67 | 4 | -4171 (-6786;-1713) | 0.094 (0.057;0.134) | 11585 (7992;16001) | 0.981 |
| 10 | 8.3 | 1.67 | 0 | -2671 (-4721;-731) | 0.073 (0.044;0.104) | 8436 (5565;11876) | 0.973 |
| 10 | 8.3 | 1.67 | -4 | -1158 (-2621;225) | 0.052 (0.031;0.075) | 5266 (3199;7789) | 0.953 |
| 10 | 8.3 | 1.67 | -8 | 325 (-548;1169) | 0.031 (0.018;0.045) | 2103 (821;3634) | 0.864 |

| | | | | | | | |
|---|---|---|---|---|---|---|---|
| 10 | 8.3 | 1.37 | 8 | -5728 (-8868;-2837) | 0.098 (0.054;0.147) | 13580 (9160;18837) | 0.98 |
| 10 | 8.3 | 1.37 | 4 | -4220 (-6784;-1838) | 0.08 (0.044;0.121) | 10609 (7003;14939) | 0.976 |
| 10 | 8.3 | 1.37 | 0 | -2683 (-4715;-836) | 0.062 (0.034;0.094) | 7684 (4807;11001) | 0.966 |
| 10 | 8.3 | 1.37 | -4 | -1197 (-2627;156) | 0.044 (0.024;0.067) | 4731 (2670;7094) | 0.946 |
| 10 | 8.3 | 1.37 | -8 | 310 (-557;1112) | 0.026 (0.014;0.04) | 1777 (502;3262) | 0.826 |
| 10 | 10 | 1.97 | 8 | -5050 (-8237;-2019) | 0.13 (0.083;0.176) | 15283 (10663;20532) | 0.985 |
| 10 | 10 | 1.97 | 4 | -3581 (-6164;-1079) | 0.106 (0.067;0.145) | 11938 (8129;16301) | 0.98 |
| 10 | 10 | 1.97 | 0 | -2088 (-4119;-154) | 0.082 (0.052;0.113) | 8548 (5626;12003) | 0.972 |
| 10 | 10 | 1.97 | -4 | -613 (-2051;802) | 0.058 (0.037;0.081) | 5184 (3081;7698) | 0.952 |
| 10 | 10 | 1.97 | -8 | 870 (-6;1723) | 0.035 (0.022;0.049) | 1887 (575;3405) | 0.829 |
| 10 | 10 | 1.67 | 8 | -5172 (-8327;-2194) | 0.115 (0.07;0.163) | 14284 (9828;19643) | 0.983 |
| 10 | 10 | 1.67 | 4 | -3667 (-6282;-1209) | 0.094 (0.057;0.134) | 11082 (7488;15497) | 0.976 |
| 10 | 10 | 1.67 | 0 | -2167 (-4217;-227) | 0.073 (0.044;0.104) | 7932 (5061;11372) | 0.969 |
| 10 | 10 | 1.67 | -4 | -655 (-2118;729) | 0.052 (0.031;0.075) | 4762 (2695;7285) | 0.944 |
| 10 | 10 | 1.67 | -8 | 829 (-44;1673) | 0.031 (0.018;0.045) | 1600 (317;3130) | 0.799 |
| 10 | 10 | 1.37 | 8 | -5225 (-8364;-2333) | 0.098 (0.054;0.147) | 13077 (8656;18333) | 0.978 |
| 10 | 10 | 1.37 | 4 | -3716 (-6280;-1334) | 0.08 (0.044;0.121) | 10105 (6499;14435) | 0.971 |
| 10 | 10 | 1.37 | 0 | -2180 (-4211;-332) | 0.062 (0.034;0.094) | 7180 (4303;10498) | 0.957 |
| 10 | 10 | 1.37 | -4 | -693 (-2123;660) | 0.044 (0.024;0.067) | 4228 (2167;6591) | 0.928 |
| 10 | 10 | 1.37 | -8 | 813 (-54;1615) | 0.026 (0.014;0.04) | 1273 (-1;2758) | 0.749 |
| 10 | 15 | 1.97 | 8 | -3569 (-6755;-537) | 0.13 (0.083;0.176) | 13802 (9181;19051) | 0.975 |
| 10 | 15 | 1.97 | 4 | -2100 (-4682;401) | 0.106 (0.067;0.145) | 10457 (6648;14819) | 0.968 |
| 10 | 15 | 1.97 | 0 | -606 (-2637;1327) | 0.082 (0.052;0.113) | 7066 (4144;10522) | 0.95 |
| 10 | 15 | 1.97 | -4 | 867 (-569;2283) | 0.058 (0.037;0.081) | 3702 (1599;6217) | 0.876 |
| 10 | 15 | 1.97 | -8 | 2351 (1475;3204) | 0.035 (0.022;0.049) | 405 (-906;1923) | 0.59 |
| 10 | 15 | 1.67 | 8 | -3690 (-6845;-712) | 0.115 (0.07;0.163) | 12802 (8346;18161) | 0.97 |
| 10 | 15 | 1.67 | 4 | -2185 (-4801;271) | 0.094 (0.057;0.134) | 9600 (6007;14015) | 0.959 |
| 10 | 15 | 1.67 | 0 | -686 (-2736;1254) | 0.073 (0.044;0.104) | 6450 (3579;9890) | 0.941 |

| | | | | | | | |
|---|---|---|---|---|---|---|---|
| 10 | 15 | 1.67 | -4 | 826 (-636;2210) | 0.052 (0.031;0.075) | 3280 (1213;5804) | 0.855 |
| 10 | 15 | 1.67 | -8 | 2311 (1437;3154) | 0.031 (0.018;0.045) | 118 (-1164;1648) | 0.532 |
| 10 | 15 | 1.37 | 8 | -3743 (-6883;-851) | 0.098 (0.054;0.147) | 11595 (7174;16851) | 0.963 |
| 10 | 15 | 1.37 | 4 | -2235 (-4798;147) | 0.08 (0.044;0.121) | 8624 (5018;12953) | 0.951 |
| 10 | 15 | 1.37 | 0 | -698 (-2729;1149) | 0.062 (0.034;0.094) | 5699 (2822;9016) | 0.92 |
| 10 | 15 | 1.37 | -4 | 787 (-642;2142) | 0.044 (0.024;0.067) | 2746 (685;5109) | 0.817 |
| 10 | 15 | 1.37 | -8 | 2295 (1427;3097) | 0.026 (0.014;0.04) | -207 (-1483;1276) | 0.464 |
| 10 | 20 | 1.97 | 8 | -2087 (-5273;944) | 0.13 (0.083;0.176) | 12320 (7699;17569) | 0.963 |
| 10 | 20 | 1.97 | 4 | -618 (-3201;1883) | 0.106 (0.067;0.145) | 8975 (5166;13338) | 0.949 |
| 10 | 20 | 1.97 | 0 | 875 (-1155;2809) | 0.082 (0.052;0.113) | 5585 (2662;9040) | 0.9 |
| 10 | 20 | 1.97 | -4 | 2349 (911;3765) | 0.058 (0.037;0.081) | 2220 (117;4735) | 0.763 |
| 10 | 20 | 1.97 | -8 | 3833 (2957;4686) | 0.035 (0.022;0.049) | -1075 (-2388;441) | 0.324 |
| 10 | 20 | 1.67 | 8 | -2208 (-5363;768) | 0.115 (0.07;0.163) | 11320 (6864;16679) | 0.954 |
| 10 | 20 | 1.67 | 4 | -704 (-3319;1753) | 0.094 (0.057;0.134) | 8118 (4525;12533) | 0.939 |
| 10 | 20 | 1.67 | 0 | 795 (-1254;2736) | 0.073 (0.044;0.104) | 4968 (2098;8409) | 0.874 |
| 10 | 20 | 1.67 | -4 | 2308 (845;3692) | 0.052 (0.031;0.075) | 1799 (-267;4322) | 0.718 |
| 10 | 20 | 1.67 | -8 | 3793 (2918;4636) | 0.031 (0.018;0.045) | -1363 (-2646;166) | 0.277 |
| 10 | 20 | 1.37 | 8 | -2261 (-5401;629) | 0.098 (0.054;0.147) | 10113 (5693;15369) | 0.943 |
| 10 | 20 | 1.37 | 4 | -753 (-3316;1628) | 0.08 (0.044;0.121) | 7142 (3536;11472) | 0.915 |
| 10 | 20 | 1.37 | 0 | 783 (-1247;2631) | 0.062 (0.034;0.094) | 4217 (1340;7534) | 0.836 |
| 10 | 20 | 1.37 | -4 | 2269 (839;3624) | 0.044 (0.024;0.067) | 1264 (-796;3627) | 0.666 |
| 10 | 20 | 1.37 | -8 | 3777 (2909;4579) | 0.026 (0.014;0.04) | -1689 (-2965;-204) | 0.229 |
| 5 | 4.3 | 1.97 | 8 | -1487 (-3181;172) | 0.082 (0.057;0.105) | 7910 (5471;10757) | 0.984 |
| 5 | 4.3 | 1.97 | 4 | -942 (-2345;420) | 0.067 (0.047;0.086) | 6203 (4198;8542) | 0.98 |
| 5 | 4.3 | 1.97 | 0 | -411 (-1505;682) | 0.052 (0.036;0.067) | 4492 (2923;6326) | 0.971 |
| 5 | 4.3 | 1.97 | -4 | 133 (-655;935) | 0.037 (0.026;0.048) | 2776 (1655;4101) | 0.95 |
| 5 | 4.3 | 1.97 | -8 | 681 (193;1170) | 0.022 (0.015;0.029) | 1069 (383;1896) | 0.846 |
| 5 | 4.3 | 1.67 | 8 | -1515 (-3130;113) | 0.075 (0.051;0.099) | 7451 (5051;10223) | 0.98 |

| | | | | | | | |
|---|---|---|---|---|---|---|---|
| 5 | 4.3 | 1.67 | 4 | -965 (-2314;357) | 0.062 (0.042;0.081) | 5839 (3863;8106) | 0.977 |
| 5 | 4.3 | 1.67 | 0 | -407 (-1476;640) | 0.048 (0.032;0.064) | 4200 (2640;5965) | 0.969 |
| 5 | 4.3 | 1.67 | -4 | 126 (-639;888) | 0.034 (0.023;0.046) | 2569 (1450;3853) | 0.934 |
| 5 | 4.3 | 1.67 | -8 | 686 (209;1152) | 0.021 (0.014;0.028) | 935 (260;1752) | 0.817 |
| 5 | 4.3 | 1.37 | 8 | -1499 (-3080;71) | 0.068 (0.044;0.091) | 6823 (4524;9561) | 0.977 |
| 5 | 4.3 | 1.37 | 4 | -952 (-2260;325) | 0.055 (0.036;0.075) | 5307 (3422;7552) | 0.97 |
| 5 | 4.3 | 1.37 | 0 | -401 (-1441;586) | 0.043 (0.028;0.058) | 3786 (2329;5536) | 0.96 |
| 5 | 4.3 | 1.37 | -4 | 142 (-618;861) | 0.031 (0.02;0.042) | 2269 (1213;3544) | 0.924 |
| 5 | 4.3 | 1.37 | -8 | 694 (225;1143) | 0.018 (0.012;0.025) | 755 (107;1553) | 0.784 |
| 5 | 8.3 | 1.97 | 8 | -302 (-1996;1358) | 0.082 (0.057;0.105) | 6725 (4286;9572) | 0.969 |
| 5 | 8.3 | 1.97 | 4 | 243 (-1160;1606) | 0.067 (0.047;0.086) | 5018 (3012;7356) | 0.953 |
| 5 | 8.3 | 1.97 | 0 | 773 (-319;1868) | 0.052 (0.036;0.067) | 3306 (1738;5140) | 0.916 |
| 5 | 8.3 | 1.97 | -4 | 1318 (530;2121) | 0.037 (0.026;0.048) | 1590 (469;2915) | 0.823 |
| 5 | 8.3 | 1.97 | -8 | 1867 (1378;2355) | 0.022 (0.015;0.029) | -115 (-802;711) | 0.454 |
| 5 | 8.3 | 1.67 | 8 | -329 (-1945;1298) | 0.075 (0.051;0.099) | 6265 (3865;9038) | 0.963 |
| 5 | 8.3 | 1.67 | 4 | 220 (-1128;1543) | 0.062 (0.042;0.081) | 4654 (2678;6921) | 0.946 |
| 5 | 8.3 | 1.67 | 0 | 777 (-290;1825) | 0.048 (0.032;0.064) | 3015 (1455;4779) | 0.901 |
| 5 | 8.3 | 1.67 | -4 | 1312 (545;2074) | 0.034 (0.023;0.046) | 1384 (265;2667) | 0.801 |
| 5 | 8.3 | 1.67 | -8 | 1872 (1394;2337) | 0.021 (0.014;0.028) | -249 (-925;566) | 0.416 |
| 5 | 8.3 | 1.37 | 8 | -314 (-1894;1256) | 0.068 (0.044;0.091) | 5637 (3339;8375) | 0.953 |
| 5 | 8.3 | 1.37 | 4 | 233 (-1074;1511) | 0.055 (0.036;0.075) | 4121 (2237;6367) | 0.929 |
| 5 | 8.3 | 1.37 | 0 | 783 (-255;1772) | 0.043 (0.028;0.058) | 2601 (1144;4351) | 0.875 |
| 5 | 8.3 | 1.37 | -4 | 1327 (566;2047) | 0.031 (0.02;0.042) | 1084 (27;2358) | 0.759 |
| 5 | 8.3 | 1.37 | -8 | 1879 (1411;2329) | 0.018 (0.012;0.025) | -429 (-1078;368) | 0.357 |
| 5 | 10 | 1.97 | 8 | 201 (-1492;1862) | 0.082 (0.057;0.105) | 6221 (3782;9068) | 0.957 |
| 5 | 10 | 1.97 | 4 | 746 (-656;2109) | 0.067 (0.047;0.086) | 4514 (2508;6853) | 0.927 |
| 5 | 10 | 1.97 | 0 | 1277 (184;2371) | 0.052 (0.036;0.067) | 2803 (1234;4637) | 0.878 |
| 5 | 10 | 1.97 | -4 | 1822 (1033;2625) | 0.037 (0.026;0.048) | 1086 (-33;2411) | 0.746 |

| | | | | | | | |
|---|---|---|---|---|---|---|---|
| 5 | 10 | 1.97 | -8 | 2371 (1882;2859) | 0.022 (0.015;0.029) | -619 (-1305;207) | 0.305 |
| 5 | 10 | 1.67 | 8 | 174 (-1441;1802) | 0.075 (0.051;0.099) | 5762 (3362;8534) | 0.95 |
| 5 | 10 | 1.67 | 4 | 723 (-624;2046) | 0.062 (0.042;0.081) | 4150 (2174;6417) | 0.922 |
| 5 | 10 | 1.67 | 0 | 1281 (212;2329) | 0.048 (0.032;0.064) | 2511 (951;4275) | 0.857 |
| 5 | 10 | 1.67 | -4 | 1816 (1049;2577) | 0.034 (0.023;0.046) | 880 (-238;2164) | 0.708 |
| 5 | 10 | 1.67 | -8 | 2375 (1898;2841) | 0.021 (0.014;0.028) | -753 (-1429;62) | 0.27 |
| 5 | 10 | 1.37 | 8 | 189 (-1390;1760) | 0.068 (0.044;0.091) | 5133 (2835;7871) | 0.93 |
| 5 | 10 | 1.37 | 4 | 736 (-571;2014) | 0.055 (0.036;0.075) | 3618 (1733;5863) | 0.894 |
| 5 | 10 | 1.37 | 0 | 1287 (248;2276) | 0.043 (0.028;0.058) | 2097 (640;3847) | 0.824 |
| 5 | 10 | 1.37 | -4 | 1831 (1070;2551) | 0.031 (0.02;0.042) | 580 (-476;1854) | 0.632 |
| 5 | 10 | 1.37 | -8 | 2383 (1914;2832) | 0.018 (0.012;0.025) | -933 (-1581;-135) | 0.209 |
| 5 | 15 | 1.97 | 8 | 1683 (-10;3343) | 0.082 (0.057;0.105) | 4739 (2300;7586) | 0.901 |
| 5 | 15 | 1.97 | 4 | 2228 (825;3591) | 0.067 (0.047;0.086) | 3032 (1027;5371) | 0.841 |
| 5 | 15 | 1.97 | 0 | 2759 (1665;3853) | 0.052 (0.036;0.067) | 1321 (-247;3155) | 0.718 |
| 5 | 15 | 1.97 | -4 | 3304 (2515;4106) | 0.037 (0.026;0.048) | -394 (-1515;930) | 0.417 |
| 5 | 15 | 1.97 | -8 | 3852 (3364;4341) | 0.022 (0.015;0.029) | -2101 (-2787;-1274) | 0.033 |
| 5 | 15 | 1.67 | 8 | 1655 (40;3284) | 0.075 (0.051;0.099) | 4280 (1880;7052) | 0.879 |
| 5 | 15 | 1.67 | 4 | 2205 (856;3528) | 0.062 (0.042;0.081) | 2668 (692;4935) | 0.815 |
| 5 | 15 | 1.67 | 0 | 2763 (1694;3811) | 0.048 (0.032;0.064) | 1029 (-530;2794) | 0.671 |
| 5 | 15 | 1.67 | -4 | 3297 (2531;4059) | 0.034 (0.023;0.046) | -601 (-1720;682) | 0.372 |
| 5 | 15 | 1.67 | -8 | 3857 (3380;4323) | 0.021 (0.014;0.028) | -2235 (-2910;-1418) | 0.024 |
| 5 | 15 | 1.37 | 8 | 1671 (90;3242) | 0.068 (0.044;0.091) | 3652 (1353;6390) | 0.849 |
| 5 | 15 | 1.37 | 4 | 2218 (910;3496) | 0.055 (0.036;0.075) | 2136 (251;4381) | 0.772 |
| 5 | 15 | 1.37 | 0 | 2769 (1729;3757) | 0.043 (0.028;0.058) | 615 (-841;2365) | 0.6 |
| 5 | 15 | 1.37 | -4 | 3313 (2552;4032) | 0.031 (0.02;0.042) | -901 (-1957;373) | 0.318 |
| 5 | 15 | 1.37 | -8 | 3865 (3396;4314) | 0.018 (0.012;0.025) | -2415 (-3063;-1617) | 0.012 |
| 5 | 20 | 1.97 | 8 | 3165 (1471;4825) | 0.082 (0.057;0.105) | 3258 (818;6104) | 0.815 |
| 5 | 20 | 1.97 | 4 | 3710 (2306;5073) | 0.067 (0.047;0.086) | 1550 (-454;3889) | 0.697 |

| | | | | | | | |
|---|---|---|---|---|---|---|---|
| 5 | 20 | 1.97 | 0 | 4241 (3147;5335) | 0.052 (0.036;0.067) | -160 (-1729;1673) | 0.469 |
| 5 | 20 | 1.97 | -4 | 4785 (3997;5588) | 0.037 (0.026;0.048) | -1876 (-2997;-551) | 0.169 |
| 5 | 20 | 1.97 | -8 | 5334 (4846;5823) | 0.022 (0.015;0.029) | -3583 (-4269;-2755) | 0.001 |
| 5 | 20 | 1.67 | 8 | 3137 (1521;4766) | 0.075 (0.051;0.099) | 2798 (398;5570) | 0.786 |
| 5 | 20 | 1.67 | 4 | 3687 (2338;5010) | 0.062 (0.042;0.081) | 1186 (-789;3453) | 0.644 |
| 5 | 20 | 1.67 | 0 | 4244 (3176;5292) | 0.048 (0.032;0.064) | -451 (-2012;1312) | 0.42 |
| 5 | 20 | 1.67 | -4 | 4779 (4012;5541) | 0.034 (0.023;0.046) | -2082 (-3202;-799) | 0.147 |
| 5 | 20 | 1.67 | -8 | 5339 (4861;5805) | 0.021 (0.014;0.028) | -3716 (-4392;-2900) | 0.001 |
| 5 | 20 | 1.37 | 8 | 3153 (1572;4724) | 0.068 (0.044;0.091) | 2170 (-128;4908) | 0.738 |
| 5 | 20 | 1.37 | 4 | 3700 (2392;4978) | 0.055 (0.036;0.075) | 654 (-1229;2899) | 0.585 |
| 5 | 20 | 1.37 | 0 | 4250 (3211;5239) | 0.043 (0.028;0.058) | -866 (-2322;883) | 0.364 |
| 5 | 20 | 1.37 | -4 | 4795 (4033;5514) | 0.031 (0.02;0.042) | -2383 (-3439;-1108) | 0.097 |
| 5 | 20 | 1.37 | -8 | 5347 (4878;5796) | 0.018 (0.012;0.025) | -3896 (-4545;-3099) | 0 |

**Table S1. Extended results for different scenarios.** An extension of table 3. Results are presented per patient with an LVO. Probabilistic sensitivity results for different scenarios. From top to bottom, the scenarios become less favorable for the NCCT+CTA+CTP compared to NCCT+CTA based large vessel occlusion detection (LVO). All scenarios considered an adjusted common OR of 1.67 for the treatment effect of EVT (12). The sensitivity difference of additional CTP represents the percentage-point difference between baseline values of 8% for ICA and 16% for M1 and M2 occlusions. NMB: Net monetary benefit at a willingness to pay of €80,000 per QALY. Δ: the difference between tNCCT+CTA+CTP and NCCT+CTA based LVO detection. ΔCosts: difference in costs; negative values imply a cost saving for the NCCT+CTA+CTP arm compared to the NCCT+CTA arm. ΔQALYs: difference in quality-adjusted life years; positive values imply a health gain for the NCC+CTA+CTP arm compared to the NCCT+CTA arm. NNI: number needed to image before one EVT eligible LVO is detected.

**Supplementary material references**

10. Jansen IGH, Mulder MJHL, Goldhoorn RJB. Endovascular treatment for acute ischaemic stroke in routine clinical practice: Prospective, observational cohort study (MR CLEAN Registry). BMJ. 2018;360.

11. Van Voorst H, Kunz WG, Van Den Berg LA, Kappelhof M, Pinckaers FME, Goyal M, et al. Quantified health and cost effects of faster endovascular treatment for large vessel ischemic stroke patients in the Netherlands. J Neurointerv Surg. 2020;1–8.

12. Berkhemer OA, Fransen PSS, Beumer D, van den Berg LA, Lingsma HF, Yoo AJ, et al. A Randomized Trial of Intraarterial Treatment for Acute Ischemic Stroke. N Engl J Med. 2015;372(1):11–20.
